# The spatio-temporal landscape of lung pathology in SARS-CoV-2 infection

**DOI:** 10.1101/2020.10.26.20219584

**Authors:** André Figueiredo Rendeiro, Hiranmayi Ravichandran, Yaron Bram, Steven Salvatore, Alain Borczuk, Olivier Elemento, Robert Edward Schwartz

**Author notes:** Co-first authors (André Figueiredo Rendeiro, Hiranmayi Ravichandran). Co-senior authors (Alain Borczuk, Olivier Elemento, Robert Edward Schwartz).

## Abstract

Recent studies have provided insights into the pathology and immune response to coronavirus disease 2019 (COVID-19)^1–8^. However thorough interrogation of the interplay between infected cells and the immune system at sites of infection is lacking. We use high parameter imaging mass cytometry^9^ targeting the expression of 36 proteins, to investigate at single cell resolution, the cellular composition and spatial architecture of human acute lung injury including SARS-CoV-2. This spatially resolved, single-cell data unravels the disordered structure of the infected and injured lung alongside the distribution of extensive immune infiltration. Neutrophil and macrophage infiltration are hallmarks of bacterial pneumonia and COVID-19, respectively. We provide evidence that SARS-CoV-2 infects predominantly alveolar epithelial cells and induces a localized hyper-inflammatory cell state associated with lung damage. By leveraging the temporal range of COVID-19 severe fatal disease in relation to the time of symptom onset, we observe increased macrophage extravasation, mesenchymal cells, and fibroblasts abundance concomitant with increased proximity between these cell types as the disease progresses, possibly as an attempt to repair the damaged lung tissue. This spatially resolved single-cell data allowed us to develop a biologically interpretable landscape of lung pathology from a structural, immunological and clinical standpoint. This spatial single-cell landscape enabled the pathophysiological characterization of the human lung from its macroscopic presentation to the single-cell, providing an important basis for the understanding of COVID-19, and lung pathology in general.

## Main text

Severe acute respiratory syndrome coronavirus-2 (SARS-CoV-2) is a novel coronavirus which has spread to become a global pandemic where over 40 million have been infected, resulting in over 1 million fatalities as of October 2020^10,11^. A growing body of evidence indicates that the severity of coronavirus disease 2019 (COVID-19) is driven by an inflammatory syndrome caused by hyper-activation of the immune system^8,12^ in an attempt to clear the virus. Persistent inflammation can result in damage to lung tissue^13^, exudation of pulmonary-edema fluid leading to dyspnea and acute respiratory distress syndrome^14,15^ (ARDS). Immune profiling in peripheral blood^1–3,5,7,16^ or bronchoalveolar lavage fluid^17^ have brought to light major changes in the immune system such as excessive neutrophil activation^18^, lymphopenia^3^, and aberrant adaptive immune system responses^2^ among the most prominent changes. However, thorough analysis of infected tissue and immune system in a spatial context has only recently started^4,19–21^ and is currently lacking for most infected organs, including the lung. While most COVID-19 patients with severe disease develop ARDS, administering routine clinical supportive care as for other ARDS does not entirely aid in patient recovery. Thus, it is unclear to what degree SARS-CoV-2 infection and immune response to COVID-19 resembles or differs from other insults in the lung. To elucidate the cellular composition, spatial context, and interplay between immune and structural cell types during SARS-CoV-2 infection in the lung, we performed imaging mass cytometry in post-mortem lung tissue from patients with COVID-19, other ARDS-causing lung infections, and otherwise healthy individuals.

### Pathophysiology of SARS-CoV2 infection in the lung

We investigated a cohort of 23 patients who died with post-influenza ARDS (n = 2), ARDS post bacterial infection (n = 4), acute bacterial pneumonia (n = 3), COVID-19 respiratory distress syndrome (n = 10), and otherwise healthy individuals (n = 4) from which post-mortem lung tissue was available (**Figure 1a, Extended Data 1, Supplementary Table 1**). In order to better understand and capture anatomic manifestations of lung disease progression, COVID-19 patients were divided into “early” and “late” disease, dependent on whether death occurred 14 days before or after 30 days from the start of respiratory symptoms, respectively (**Supplementary Table 1**). To comprehensively investigate the cellular environment and spatial organization of the lung, we designed an imaging mass cytometry (IMC) based metal-labeled antibody panel composed of 36 biomarkers, and used it to generate 237 highly multiplexed images at 1 µm resolution, totaling 332 mm^2^ of tissue profiled and 664,006 single cells identified across all specimens. IMC leverages inductively coupled plasma mass spectrometry (**ICP**-**MS**) based laser ablation of lanthanide metal-tagged antibodies from tissues for the quantitative detection of epitope abundance in a spatially-resolved manner (**Figure 1a** and **Supplementary Table 2**). Our panel included phenotypic markers of endothelial, epithelial, mesenchymal, and immune cells, functional markers (activation, inflammation and cell death), as well as an antibody specific to the Spike protein of SARS-CoV-2. Driven by the observation that post-mortem patient lungs showed a considerable increase in weight across all pathologies **(Figure 1b)**, we employed the IMC data in order to quantify the histopathology of the lung under infection (**Methods**). Consistent with the gain of weight during infection, the lacunar space of infected lungs was significantly reduced from 41.1% in the healthy lung to median ranges of 28.72% and 15.3% in Flu and 15.3% in “late” COVID-19, respectively (**Figure 1c**,**e**) with the most pronounced change being associated with the alveolar epithelium **(Extended Data 2a**,**b**). Since collagen deposition is a known mediator of both normal and dysregulated tissue repair during infection recovery^22^, we quantified the extent and intensity of Collagen deposition into a unified fibrosis score, in a manner inspired by the Ashcroft score of pulmonary fibrosis^23^ (**Methods**). The fibrosis score was significantly higher for all lung pathologies when compared with the normal lung, especially in COVID-19 (**Figure 1d**,**f, Extended Data 2c-e**). In order to understand the cellular composition of the lung during various lung insults, we constructed a spatially resolved single-cell atlas by segmenting images into single-cells, and quantified the abundance of each marker in each cell (**Methods**). The 664,006 single-cells detected across all disease groups were projected into a two-dimensional space using the Uniform Manifold and Projection (UMAP) algorithm (**Figure 1g** and **Extended Data 3a**) and clustered based on their phenotype in an unsupervised manner using graph clustering (**Figure 1h** and **Extended Data 3b**), resulting in a single-cell phenotypic atlas for the human lung. We identified a total of 36 cellular clusters, which we organized into 17 meta-clusters based on the predominant markers, overall phenotypes, and proximity to vessel, airway and alveolar lacunae (**Figure 1i** and **Extended Data 3b, 4**). This atlas was dominated by abundant structural cell types such as Keratin 8/18^+^ alveolar epithelial cells, α-smooth muscle actin^+^ (α-SMA) cells lining vasculature, and immune cells such as CD15^+^, CD11b^+^ polymorphonuclear neutrophils or CD68^+^ macrophages (**Figure 1j**). While the broad compartments of lung structural cells and immune cells did not present with large changes in absolute

**Figure 1:**
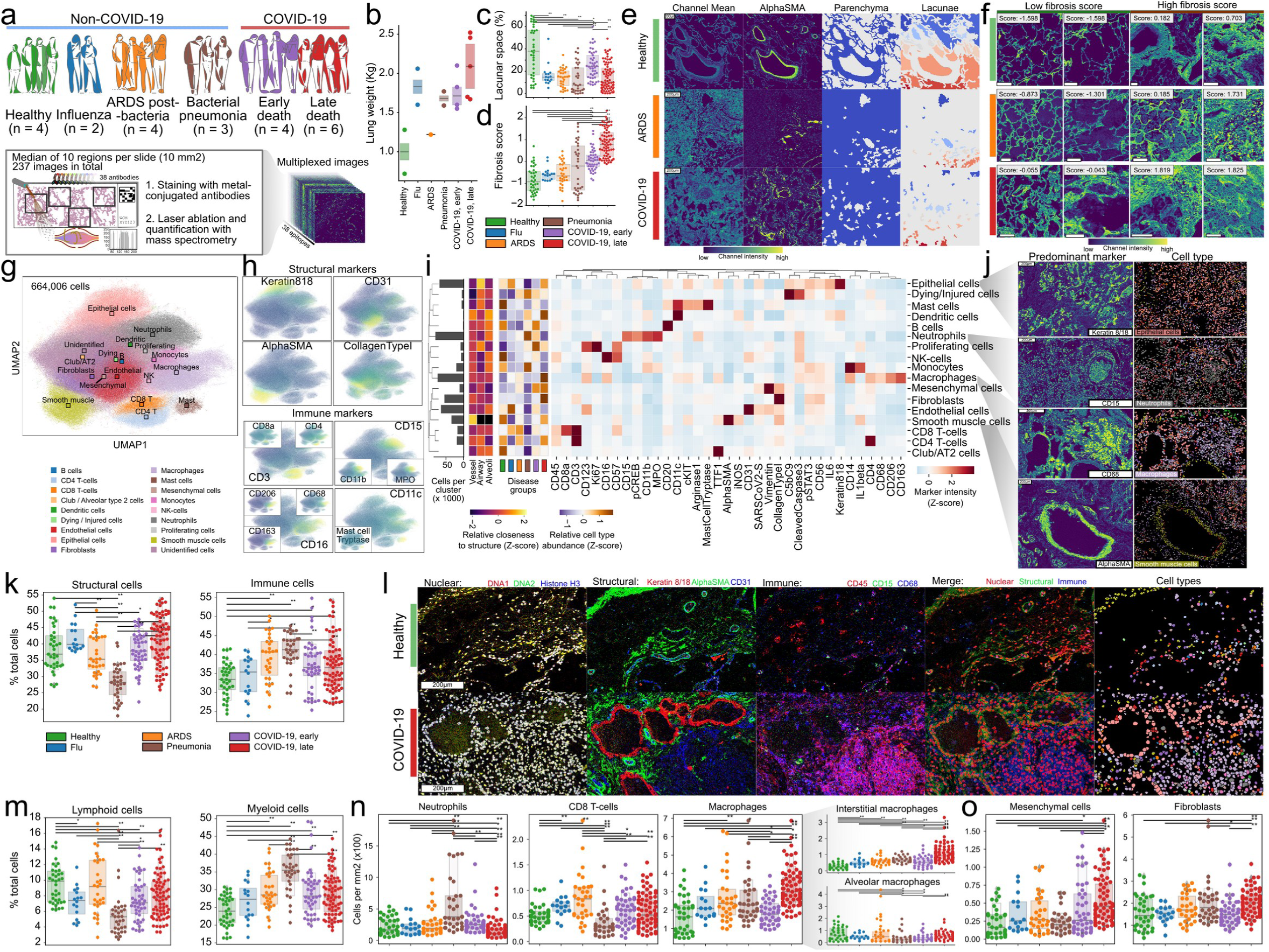
Structural and immunological disorder of lung infection. **a)** Composition of lung infection cohort and schematic procedure to acquire highly multiplexed spatially resolved data with imaging mass cytometry (IMC) from post-mortem lung samples. **b)** Total lung weight per disease group measure at autopsy. **c)** Lacunar space for each acquired IMC image as percentage of image area. **d)** Fibrosis score for each acquired IMC image. **e)** Representative images illustrating the lacunar and parenchymal structure of lungs from healthy individuals, patients with ARDS or COVID-19. The mean of all acquired channels illustrates the general tissue structure, while α-SMA the vasculature of the tissue. **f)** Collagen type I in images from lungs of healthy individuals, patients with ARDS or COVID-19 and the associated fibrosis score. Images with lowest and highest fibrosis scores are depicted. **g)** UMAP representation of all quantified cells and the meta-cluster label associated with each. Meta-cluster centroids are represented with squares. **h)** UMAP representation as in g) but with color scale representing the intensity of each marker in each single-cell. **i)** Phenotype of each meta-cluster characterized by the average intensity of each marker. The histogram indicates the abundance of each meta-cluster. The heatmaps on the left indicate the relative proximity of each meta-cluster to lung structures or abundance in each disease group. **j)** Representative spatial context of three meta-clusters (rows). The column on the left displays the spatial distribution of the most predominant marker for each meta-cluster, while the column on the right represents segmented cells colored by the meta-cluster they were assigned to. **k)** Relative abundance of structural or immune cells in images of each disease group. **l)** Representative images of the spatial distribution of structural and immune cells in lungs from a healthy individual and a COVID-19 patient. **m)** Relative abundance of lymphoid or myeloid cells of the immune system for each disease group. **n-o)** Absolute abundance of n) immune or o) structural cell types as identified by the corresponding meta-cluster for images in each disease group. ** p < 0.01; * p < 0.05, Mann-Whitney U-test, pairwise between groups, Benjamini-Hochberg FDR adjustment. numbers (**Extended Data 5**), the specific internal composition of the structural cells of the lung and the immune system changed extensively (**Figure 1k**). We observed increased immune infiltration in COVID-19 when compared with healthy lung, but to a degree comparable with other lung infections (**Figure 1k**,**l** and **Extended Data 5**). Within the specific immune components, we observed a prominent increase in infiltration of myeloid cells in COVID-19 compared with healthy lung, but to a lesser extent than in pneumonia (**Figure 1m, Extended Data 6-7**). More detailed examination of the phenotypic diversity of myeloid cells in respect to their location in the lung (**Extended Data 8**), revealed that CD14^+^, CD16^+^, CD206^+^, CD163^+^, CD123^+^ interstitial macrophages, a population likely recruited from peripheral blood displayed the greatest increase in COVID-19 when compared with healthy lung, particularly in late disease (**Figure 1n**), and the highest expression of IL1β in monocytes in early disease (**Extended Data 8**). While in early COVID-19 neutrophils are abundant to a similar level as in healthy lungs, they are present in significantly lower absolute amounts in late disease (**Figure 1n**, and **Extended Data 6-7**); a stark contrast with pneumonia which harbors the highest amount of neutrophils across all disease groups. Lung macrophage populations were particularly increased in COVID-19 compared to all other disease states (**Figure 1n, Extended Data 6-7**). We also observed that CD8^+^ T cells were significantly increased in non-COVID-19 ARDS, but depleted in pneumonia in comparison with the healthy lung (**Figure 1n, Extended Data 6-7**). Specifically in SARS-CoV-2 infected lungs, while the abundance of alveolar epithelial and smooth muscle cells was comparable with healthy lungs, we observed a shift in the stromal compartment of the lung, with a significant reduction in absolute numbers of endothelial cells and an increase of mesenchymal cells and fibroblasts in late COVID-19 (**Figure 1o, Extended Data 6-7**). The increase in fibroblast abundance in COVID-19 is consistent with the previously discussed and observed advanced fibrosis in COVID-19 lung (**Figure 1d, Extended Data 7c**).

### Widespread inflammation in SARS-CoV-2^+^ lungs leads to tissue damage

The established lung phenotypic single-cell atlas contains clusters defined by cell identity markers and markers of cell state across all diseases. As a complementary approach, we developed unsupervised classification of cells according to the abundance of each marker (**Extended Data 9a**), observing high concordance with the phenotypic clusters (**Extended Data 9b**). Using this approach, we observed high specificity of our SARS-CoV-2 Spike antibody to tissue samples from COVID-19 patients (**Figure 2a** and **Extended Data 9c**). Among all cell types, alveolar epithelial cells displayed the highest rate of SARS-CoV-2 Spike (SARS-CoV-2 Spike^+^) positivity (**Figure 2b**). These alveolar epithelial cells were also highly positive for the phosphorylated signal transducer and activator of transcription 3 (pSTAT3), the receptor tyrosine kinase and proto-oncogene c-KIT, and contained increased levels of interleukin 6 (IL-6), Arginase1, the apoptosis marker cleaved Caspase 3, and the assembled Complement membrane attack complex C5b-C9 (**Figure 2c**). Given that SARS-CoV-2 Spike-negative alveolar epithelial cells in the same regions showed no increase in the levels of functional markers, this likely indicates viral-specific alterations of the cellular state. Increased IL-6 and pSTAT3 levels indicating an inflammatory state were also elevated in Flu and pneumonia when compared with healthy lung but not in non-COVID-19 ARDS (**Extended Data 9d**). However, high levels of cleaved Caspase3 and C5b-C9 were exclusive to COVID-19 SARS-CoV-2 Spike+ alveolar epithelial cells indicating initiation of apoptosis and complement-mediated host immune defense, respectively, leading to increased alveolar lining damage. While the alveolar epithelium was the predominant cell type positive for SARS-CoV-2 Spike protein, we also found that a mean of 7.8% and 2.7% of macrophages and neutrophils were also positive for SARS-CoV-2 Spike protein respectively across all COVID-19 images, with some regions with up to 38.6% and 43.6% positive respectively (**Extended Data 9e-g**). Consistent with the observation in SARS-CoV-2 Spike+ epithelial cells, both macrophages and neutrophils exhibited higher levels of cleaved Caspase3, pSTAT3 and IL-6, but unlike the alveolar epithelial cells, show no C5b-C9 positive staining (**Extended Data 12**). cKIT however, was specifically upregulated in macrophages and not in neutrophils (**Extended Data 10**). This inflammatory phenotype seen in SARS-CoV-2 Spike^+^ cells was also seen in other cell types, albeit at a much lower frequency (**Extended Data 11**). Despite the pervasive inflammatory phenotype of SARS-CoV-2 Spike^+^ cells, we observed high heterogeneity in the localization of these cells, often within the same 1 mm^2^ tissue region (**Figure 2e**). While clear interactions between SARS-CoV-2 Spike^+^ epithelial cells and immune and non-immune cells could be observed (**Figure 2e, upper right**), other SARS-CoV-2 Spike^+^ cells did not interact with these cells at all, or seemed encapsulated in structures which precluded interactions with other cell types (**Figure 2e, lower right**). In order to generate a quantitative map of cellular interactions, we quantified proximal interactions between and within cell types for each image, and generated disease-specific interaction maps (**Figure 2f, Extended Data 12** and **Methods**). This revealed cell types such as B cells which preferentially interact with themselves, CD8^+^ T cells that have very limited interactions at all, and pervasive interactions between structural and immune cells across all diseases (**Figure 2f**). Comparing interactions between the healthy lung and COVID-19, we observed increased interactions between neutrophils and macrophages and decreased interactions within macrophages, while in late COVID-19 disease the intra-cell type interactions in macrophages, fibroblasts and CD4^+^ T cells were further decreased, while accompanied by an increase in interactions between macrophages and fibroblasts or dendritic cells (**Figure 2h**,**i**). When spatially contextualizing these interactions, we observed that macrophages preferentially interact with fibroblasts in the alveolar walls, suggesting a contribution to fibrosis and thickening of the alveolar wall in late disease (**Figure 2j**). Since epithelial cells as a whole did not show any particular change in interactions in COVID-19 compared with healthy lung, we investigated whether SARS-CoV-2 Spike^+^ epithelial cells differed in cellular interactions to their SARS-CoV-2 Spike^-^ counterparts in COVID-19 lungs (**Extended Data 14**). Across all cell types, there was a trend for SARS-CoV-2 Spike^+^ cells to have reduced cellular interactions with other cell types, regardless of whether these were also SARS-CoV-2 Spike^+^ (**Extended Data 13a-c**). SARS-CoV-2 Spike^+^ epithelial cells in particular lacked interactions with other cell types when compared with SARS-CoV-2 Spike-cells (**Extended Data 13d-f**). Perhaps as the consequence of the lack of specific interactions between immune cells and infected cells, we observed progressively more cells with the markers of cell death, with a preference for immune cells such as macrophages and neutrophils to have the apoptosis marker Cleaved Caspase3 (**Figure 2k**), while structural cell types such as epithelial cells and fibroblasts were preferentially marked with C5b-C9 (**Figure 2l**).

**Figure 2:**
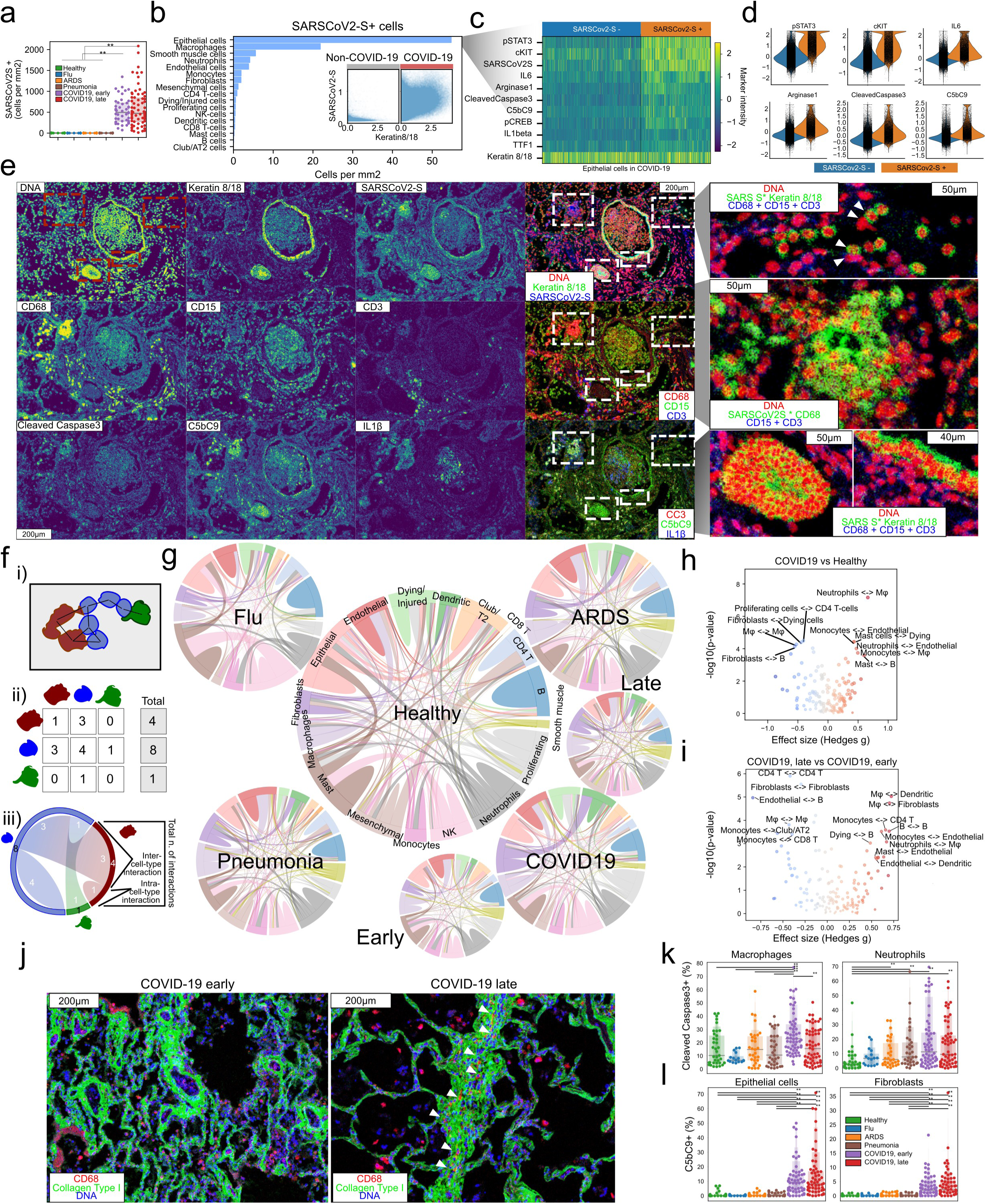
Cellular tropism of SARS-CoV-2 infection. **a)** Absolute abundance of SARS-CoV-2 Spike^+^ cells in each image for each disease group. **b)** Distribution of SARS-CoV-2 Spike^+^ cells across all meta-clusters of COVID-19 cells. The inset figure displays the intensity of Keratin 8/18 and SARS-CoV-2 Spike^+^ for each single cell. **c)** Phenotypic profile of alveolar epithelial cells depending on SARS-CoV-2 Spike levels. **d)** Intensity levels per single-cell of differential markers between cells dependent on SARS-CoV-2 Spike levels. **e)** Image illustrative of the distribution of SARS-CoV-2 Spike signal in a spatial context. Structural, cell type specific and functional markers are displayed either alone or in combination. The rightmost column contains zoomed in images of the highlighted regions. For the green channel in the rightmost column images, the SARS-CoV-2 Spike channel was multiplied with either Keratin 8/18 or CD68 channel in order to highlight cells positive for both markers, since the images were scaled to the unit interval. **f)** Schematic depiction of the procedure to quantify and represent cellular interactions between cell types. i) a graph is built connecting nearby cells with edges; ii) the edges between each cell type are quantified in a pairwise manner; iii) A chord plot where cellular interactions are displayed as a proportion of all interactions found, and edges connecting cell types represent more abundant interactions. More details in Methods. **g)** Chord plots illustrating disease type-specific cellular interactions between meta-clusters in the human lung. **h-i)** Differential interactions between h) Healthy lung and COVID-19, or i) COVID-19 early and COVID-19 late. The y-axis represents the significance in interaction change [-log10(p-value)] in a two-tailed Mann-Whitney test, and the x-axis an effect size as given by a Hedges’g measure. **j)** Representative images containing fibroblasts and macrophages from COVID-19 early and late, in which these two cell types significantly interact more in late COVID-19. **k-l)** Proportion of k) cleaved Caspase 3^+^ or l) C5b-C9^+^ cells as a percentage of the number of cells of the same cell type for each disease group. **p < 0.01; *p < 0.05, Mann-Whitney U-test, pairwise between groups, Benjamini-Hochberg FDR adjustment.

### A biologically interpretable, clinically annotated landscape of lung infection

Building on the identified cell types, their functional status, and their interactions, we sought to define a landscape of lung pathology from the data in order to form an unbiased view of the multicellular architecture of the tissue during infection (**Figure 3a** and **Extended Data 14-15** and **Methods**). The major axes of this Principal Component Analysis (PCA)-based landscape demonstrate at the same time the distribution of samples, and the major drivers of the establishment of the space. This confers biological interpretability to the landscape because the underlying cellular composition at each given point is readily identifiable. While determined in an unsupervised manner (without knowledge of sample groups), this space largely recapitulates the disease ontogeny of the samples, being dominated by the difference between samples of healthy lung to lungs of COVID-19 patients who succumbed after prolonged disease (**Figure 3a**). In order to exemplify the various phenotypes across the landscape through the images that gave rise to it, we annotate the PCA space with the images closest to the edges of the space and therefore most representative of the area (**Figure 3a**). Healthy lung samples occupy a space characterized by SARS-CoV-2 Spike^-^ alveolar epithelial and vascular endothelial cells, along with B cells, Mast cells, Monocytes and NK-cells, all interacting in intact alveolar and vascular space. Samples of COVID-19 patients with late disease occupied the opposite end of the landscape, which is characterized by SARS-CoV-2 Spike^+^ alveolar epithelial cells, CD163^+^, CD206^+^ macrophages, Collagen^+^ fibroblasts and, Vimentin^+^ mesenchymal cells, often with thick alveolar walls or even with undefined or disrupted alveolar structure. Both Influenza and ARDS groups are distinct from lungs of healthy individuals or patients with late COVID-19 disease and lie at the center of the landscape, while samples from early COVID-19 disease lungs are heterogenous and spread across the spectrum between healthy and late COVID-19, often displaying higher neutrophil infiltration within the largely lacunar structure of the tissue than late COVID-19. Lungs from patients with pneumonia share the heterogeneous profile with early COVID-19. A group of samples occupies an area characterized by the highest neutrophil infiltration across all diseases and extensive monocyte populations, while other samples are positioned more in the center of the landscape. Of note, the structure of the landscape was largely recapitulated with complementary methods (**Extended Data 15**), indicating the underlying structure revealed by the landscape is indifferent to the choice of algorithm and likely reflecting fundamental principles in the multicellular architecture of the lung. While the landscape is intrinsically biologically linked, we sought to further augment it with a layer of clinical interpretability. To that end, we performed an association analysis between the landscape axes (PCA components) and known factors of demographic, temporal and clinical nature of lung infection patients (**Figure 3b, Extended Data 16**, and **Methods**). We observed strong associations primarily between clinical factors and the first principal component (**Extended Data 16**), namely significant positive association between the number of days elapsed since beginning of symptoms and hospitalization and intubation, lung weight at death, and patient temperature at admission and the distribution of samples in the first PC axis (**Figure 3b** and **Extended Data 16**). We also observed a significant association between the reduction in white blood cell counts (WBC) and the major axis associated with disease progression (PC 1) (**Figure 3b** and **Extended Data 16**). The values of clinical factors, when overlaid with their respective images in the landscape, confer a convenient way of interpreting the discovered associations, effectively rendering a landscape annotated with clinical information, and both biologically (**Figure 3a**) and clinically interpretable (**Figure 3c** and **Extended Data 17**).

**Figure 3:**
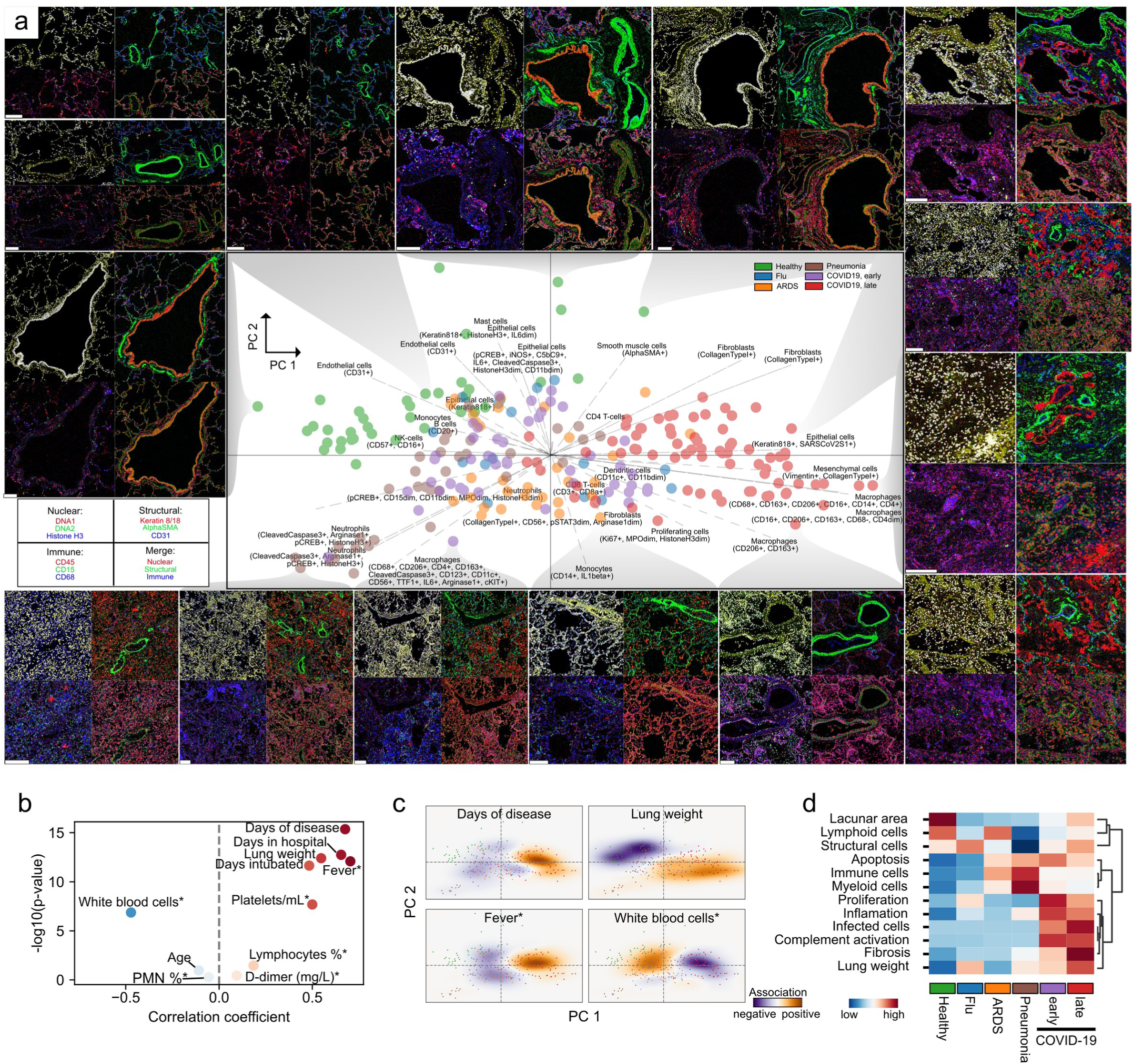
A data-driven, clinically annotated landscape of lung pathology. **a)** (center) Principal Component Analysis of IMC images from healthy and infected lungs. Points representing images are coloured by disease group. Arrows pointing outward from origin are vectors for each cluster. The vector direction indicates the area where each cell type is most abundant, while the length of the vector indicates how much it contributes to the establishment of the landscape. (contour) Image quadruplets illustrate the image closest to the border of the PCA plot at that point A white scale bar represents 200 microns. **b)** Volcano plot showing the direction and strength (x-axis) of the association between clinical parameters and Principal Component 1 (PC 1) (x-axis), as well as the significance of the association (y-axis). Significance was assessed with an empirical p-value over 10^6^ permutations (Methods). An asterisk indicates the clinical parameter was measured at admission. **c)** Projection of significantly associated clinical parameters onto the two-dimensional PCA space. The background color represents the difference between kernel density functions fitted with and without the clinical parameter as weight. An asterisk indicates the clinical parameter was measured at admission. **d)** Summary of major measured factors and their distribution across disease types. Colour scale represents a row-wise Z-score.

## Discussion

Our spatially resolved single-cell analyses of post-mortem lung tissue from COVID-19 patients and other lung infections provides a comprehensive examination of the response of the human lung to infection from the macroscopic to the single-cell level (**Figure 3d**). Across all diseases, we observed a significant reduction in alveolar lacunar space, increased immune infiltration, and cell death by apoptosis compared with healthy lungs. However, we also noted that neutrophil infiltration, while increased in ARDS and early COVID-19 compared with normal lung, is a hallmark of bacterial pneumonia, and that a high degree of inflammation, infiltration of interstitial macrophages, complement activation and fibrosis is particular to late COVID-19. While our analysis agrees with recent reports that indicate the type of pathophysiological response to SARS-CoV-2 infection may not be entirely different from ARDS unrelated to COVID-19^14^, it is at odds with reports suggesting that the hyper-inflammatory phenotype as assessed by cytokine levels in peripheral blood is not COVID-19 specific^15^. Our observation that SARS-CoV-2 Spike^+^ alveolar epithelial cells do not differentially interact with cells of the immune system despite extensive immune infiltration in the lung, potentially highlights the lack of “on-target” immunological response, while the high amount of complement activation in COVID-19 lung tissue likely results in indiscriminate “off-target” tissue damage, exacerbating COVID-19 disease and continuing the cycle of inflammation. While the observed increased presence of IL-1β+ monocytes in early disease suggests a mechanism for neutrophil recruitment to the lung, their contribution to disease pathology — and in particular that of neutrophil extracellular traps (NETs) — remains to be elucidated. Likely in response to the extensive tissue damage, late COVID-19 disease specifically displays hallmarks of tissue healing such as the expansion of mesenchymal cells and fibroblasts, possibly favoured by increased interactions with interstitial macrophages. However, the high mortality rate of COVID-19 is at odds with productive recovery from tissue damage and healing, suggesting complement-activation induced damage to the lung together with additional immunological factors such as NETs could result in microthrombi formation^19^. This raises the possibility that early immunological interventions that diminish the immunologic response and/or target the effectors of the complement cascade could have therapeutic benefit. Our biologically interpretable, clinically annotated landscape of lung pathology provides a framework for data-driven, spatially aware understanding of lung pathology, and an important resource for the study of COVID-19 and other lung infections.

## Methods

### Data reporting

No statistical methods were used to predetermine sample size. Image acquisition, segmentation, quantification, and clustering were blinded to patient identifiers and clinical metadata.

### Human studies

Tissue samples were provided by the Weill Cornell Medicine Department of Pathology. The Tissue Procurement Facility operates under Institutional Review Board (IRB) approved protocol and follows guidelines set by Health Insurance Portability and Accountability Act (HIPAA). Experiments using samples from human subjects were conducted in accordance with local regulations and with the approval of the IRB at the Weill Cornell Medicine. The autopsy samples were collected under protocol 20-04021814.

### Tissue section preparation

Lung tissues were fixed via 10% neutral buffered formalin inflation, sectioned and fixed for 24 hours prior to processing and embedding into paraffin blocks. Freshly cut five micron sections were mounted onto charged slides.

### Antibody panel design and validation

We designed an antibody panel capturing different immune and stromal compartments of the lung. Antibody clones were extensively validated through immunofluorescence and chromogenic staining and verified by a pathologist. Once the clone was approved, 100 µg of purified antibody in BSA and Azide free format was procured and conjugated using the MaxPar X8 multimetal labeling kit (Fluidigm) as per the manufacturer’s protocol. In order to confirm the antibody binding specificity post conjugation and to identify the optimal dilution for each custom conjugated antibody, sections from healthy, pneumonia ARDS infected, and SARS-CoV-2 infected lung were stained. These sections were then ablated on Fluidigm Hyperion Imaging System and visualized using MCD Viewer for expected staining pattern and optimal dilution that presented with good signal-to-noise ratio for each channel. For channels with visible spill-over into the neighboring channels, a higher dilution factor was adopted while staining the cohort tissues.

### Imaging mass cytometry

Based on the clinico-pathological characteristics and quality of the preserved tissues, suitable representative fresh cut 4-micron thick FFPE sections were acquired from the Pathology department for IMC staining. The tissues were stored at 4°C for a day before staining. Slides were first incubated for 1 hour at 60°C on a slide warmer followed by dewaxing in fresh CitriSolv (Decon Labs) twice for 10 minutes, rehydrated in descending series of 100%, 95%, 80%, and 75% ethanol for 5 minutes each. After 5 minutes of MilliQ water wash, the slides were treated with antigen retrieval solution (Tris-EDTA pH 9.2) for 30 minutes at 96°C. Slides were cooled to room temperature (RT), washed twice in TBS and blocked for 1.5 hours in SuperBlock Solution (ThermoFischer), followed by overnight incubation at 4°C with the prepared antibody cocktail containing all 36 metal-labeled antibodies (Supplementary Table 2). Next day, slides were washed twice in 0.2% Triton X-100 in PBS, and twice in TBS. DNA staining was performed using Intercalator-Iridium in PBS solution for 30 minutes in a humid chamber at room temperature. Slides were washed with MilliQ water and air dried prior to ablation.

The instrument was calibrated using a tuning slide to optimize the sensitivity of detection range. Hematoxylin and Eosin (H&E) stained slides were used to guide the selection of regions of interest containing alveolar parenchyma, airways and thrombotic vessels in order to obtain regions representative of the whole range of lung pathology. All ablations were performed with a laser frequency of 200 Hz. Tuning was performed intermittently to ensure the signal detection integrity was within the detectable range. A total of 240 image stacks were ablated. The raw .MCD files were exported for further downstream processing.

### Data preprocessing

Imaging mass cytometry data was preprocessed as previously described^24^ with some modifications. Briefly, image data was extracted from MCD files acquired with the Fluidigm Hyperion instrument. Hot pixels were removed using a fixed threshold, the image was amplified two times, Gaussian smoothing applied and, from each image a square 500-pixel crop was saved as a HDF5 file for image segmentation. Segmentation of cells in the image was performed with ilastik^25^ (version 1.3.3) by manually labeling pixels as belonging to one of three classes: nuclei – the area marked by signal in the DNA and Histone H3 channels; cytoplasm – the area immediately surrounding the nuclei and overlapping with signal in cytoplasmic channels; and background – pixels with low signal across all channels. Ilastik uses the labeled pixels to train a Random Forest classifier using features derived from the image and its derivatives. Features used were the Laplacian of the Gaussian, Gaussian Gradient Magnitude, Difference of Gaussians, Structure Tensor Eigenvalues and the Hessian of Gaussian Eigenvalues, each of them with Gaussian kernels of widths from 0.3 to 10, totaling 37 features. The outputs of prediction are class probabilities for each pixel which were used to segment the image using CellProfiler^26^ (version 3.1.8) with the *IdentifyPrimaryObjects* module. This is followed by the *IdentifySecondaryObjects* module where the identified nuclei are used to seed an expansion of the cell area to the area with the sum of the nuclear and cytoplasmatic probability map, and finally gaps in the identified cells are filled.

We assessed the quality of each acquired channel by computing a set of metrics for each channel across all images: the mean and squared coefficient of variation of each channel in the whole image and in the area with cells, a difference between those values in the cells and the whole image (foreground vs background signal), an estimate of noise variance^27^, a robust wavelet-based estimator of Gaussian noise standard deviation^27,28^, the fractal dimension (Minkowski–Bouligand approximation using the box counting method) and lacunarity of the image^29^. Across all 240 ablated images three were discarded based on these metrics and visual inspection.

### Macroscopic pathology with computer vision algorithms

In order to identify lacunae in the images we used the mean of all channels in each image stack after performing histogram equalization per channel (*skimage*.*exposure*.*equalize_hist*). Images were thresholded with Otsu’s method (*skimage*.*filters*.*threshold_otsu*), successively dilated and closed (*ski*.*morphology*.*binary_dilation/ski*.*morphology*.*closing*) with a disk of 5 um diameter in order to remove objects without holes, and for the objects with holes, objects within the hole were removed on the negative image (*scipy*.*ndimage*.*binary_fill_holes*) and, only objects with area larger than 625 pixels (25 ** 2) were kept (*skimage*.*morphology*.*remove_small_objects*). To provide biological context for the single-cell clusters identified, we further classified each lacunae of healthy lungs into three classes: blood vessels (arteries and veins), airways and alveoli. Vessels showed a very thin lining of endothelial cells, followed by a thick layer of smooth muscle cells that are α-smooth muscle actin+, while the airway epithelium is lined by Keratin 8/18+ cells, and alveoli are covered in alveolar epithelial cells that have various degrees of CD31, Vimentin and Keratin 8/18. Based on this, we developed a semi-supervised strategy for lacunae classification that had two stages: firstly each lacunae objects was dilated by a 15-pixel disk and the mean intensity of the channels above was quantified only in the dilated area and these values were Z-score transformed per image. We used these values in three ways where each providing a vote towards a lacunae being one of the three classes: absolute intensity, Z-score transformed intensity, ratio of α-smooth muscle actin Keratin 8/18. For each, a set of rules was enforced in which lacunae with higher values in α-smooth muscle actin and low in Keratin 8/18 were labeled as vessels, while higher Keratin 8/18 were labeled as airways and the remaining as alveoli. Essentially, absolute and relative intensity of the markers determine the class and the ratio of the two is the tiebreaker in case of disagreement. In a second phase, the suggested labels were reviewed by an expert and overruled if needed. In general we found the rules above accurate with only a systematic bias to underclassify vessels, and hence the needed supervision.

To develop a score for fibrosis, we were inspired by the Ashcroft score^23^ where the fraction of fibrotic tissue occupying each image is translated into a score in a Likert scale. We quantified the fraction of the image occupied by Collagen Type I as thresholded by the Otsu method (*skimage*.*filters*.*threshold_otsu*), but in addition also quantified the density of collagen per area unit by using the spectral counts given by the IMC data. The final score is the mean of a Z-score of the fraction covered by collagen and a Z-score of its intensity.

### Cell type identification

To identify cell types in an unsupervised fashion, we first quantified the intensity of each channel in each segmented cell not overlapping image borders. In addition, for each cell we computed morphological features “area”, “perimeter”, “major_axis_length”, “eccentricity”, and “solidity” (*skimage*.*measure*.*regionprops_table*). Values were Z-scored per image and cells with “area” values above −1.5, solidity above −1 and eccentricity below 1 were kept. In addition, we calculated the sum of log(1+x) signals in the IMC channels and kept cells with values between 2 and 7. We used Scanpy ^30^(version 1.6.0) to perform Principal Component Analysis, compute a neighbor graph on the PCA latent space, compute a Uniform Manifold Approximation and Projection (UMAP)^10^ embedding (*umap* package, version 0.4.6), and cluster the cells with the Leiden algorithm^31^ with resolution 1.0 (*leidenalg* package, version 0.8.1). Each cluster was manually labeled with a broad ontogeny and the channels most abundant in each cluster. These broad labels form the basis of the meta-clusters used to aggregate clusters based on cell type and regardless of cellular state. Clusters without enrichment for any particular marker were not aggregated.

To obtain an easy way to quantify the fraction of cells positive for a given marker, we employed univariate Gaussian Mixture models using scikit-learn^32^ (version 0.23.0). For each channel, we performed model selection with models with two to six mixtures, selected the model based on the Davies–Bouldin index^33^ and labeled a cell as positive for a given channel if its value was in the top mixture in case the selected model had only two mixtures or the top two mixtures if more.

### Quantification of cellular interactions

In order to quantify the degree and significance of intra- and inter-cell type interactions, we started by constructing a region adjacency graph (RAG) representing the interactions between cells, where the edges are weighted by the Euclidean distances between cells, using *scikit-image*^*34*^ (version 0.17.2). A pairwise adjacency matrix between cell clusters was computed using *networkx*^*35*^ (version 2.5). In order to get a degree of confidence on these given the cell type abundance in each image, we permuted the cell cluster assignments 1000 times and computed the difference between the log-normalized frequency of cell type interactions in the real data vs the permuted (interaction scores). For visualization, we generated chord plots by aggregating the interaction scores of the images from each disease group or subgroups by the fraction of images whose interaction score was higher than 1. To discover differential interactions specific to a subgroup, we tested whether the distribution of interaction scores between disease groups or subgroups for each pairwise cell-type combination was different as described in the next section.

### Differential testing

Differential testing was performed always pairwise between groups with a two-sided Mann-Whitney U test, and adjusted for multiple comparisons with the Benjamini-Hochberg False Discovery Rate (FDR) method using *pingouin*^*36*^ (version 0.3.7). Estimated values of central tendency, effect sizes and *p*-values are provided in tabular form in Supplementary Tables 3 and 4.

### Landscape of lung pathology and association with clinical parameters

To create the unsupervised landscape of lung pathology, we employed Principal Component Analysis (PCA) on a matrix of cell counts per cell cluster (features), per image (observations), after prior normalization by total, scaling and centering, using the *Scanpy* implementation. The feature loadings were plotted on the same dimensions as the observations by scaling them by a constant factor of 20. The correlation coefficient between each principal component (PC) and the continuous clinical variables was used as a relative measure of direction and strength of association. However the significance of association was assessed by permuting the clinical variables 10^6^ times and using the mean and standard deviation of the correlation coefficients from the permuted data as location and scale parameters respectively of a normal distribution from which the 2 * CDF(|x|) was calculated as a two-tailed empirical p-value. All principal components and clinical factors were permuted and the empirical p-values were adjusted with the Benjamini-Hochberg FDR method. We used the signed empirical, FDR corrected p-values as an effectively regularized measure of association between PCs and clinical factors. To project the clinical parameters into the PCA space, we fit Gaussian kernel density estimator functions to the distribution of the images in the first principal components, in one case without and another with the numeric values of the clinical variables as weights. The difference in predicted densities in the two-dimensional space between the weighted and unweighted kernel was used as a visual aid to identify regions in the latent space with relatively higher or lower fraction of samples with that parameter.

Additional software versions used: Python version 3.8.2, numpy^37^ version 1.18.5, scipy^38^ version 1.4.1 and scikit-image^34^ 0.17.2, *networkx*^*35*^ version 2.5, Scanpy version 1.6.0, *pingouin*^*36*^ version 0.3.7.

### Data availability

Raw IMC data are available at the following URL: https://doi.org/10.5281/zenodo.4110560

### Code availability

The source code for data preprocessing and analysis is available at the following URL: https://github.com/ElementoLab/covid-imc

## Data Availability

Raw imaging mass cytometry data are available at a Zenodo repository: https://doi.org/10.5281/zenodo.4110560
Source code for data preprocessing and analysis is available at a GitHub repository: https://github.com/ElementoLab/covid-imc

https://github.com/ElementoLab/covid-imc

## Acknowledgements

We honor the patients in this study who succumbed to disease, and the sacrifice of all physicians, nurses and allied health workers fighting the COVID-19 pandemic. We apologize to the colleagues which work we could not cite due to editorial constraints. This work was partially supported by the WorldQuant Initiative for Quantitative Prediction. A.R. is supported by a NCI T32CA203702 grant. O.E. is supported by Volastra, Janssen and Eli Lilly research grants, NIH grants UL1TR002384, R01CA194547, and Leukemia and Lymphoma Society SCOR 7012-16, SCOR 7021-20 and SCOR 180078-02 grants. R.E.S is supported by NIH grants NCI R01CA234614, NIAID 2R01AI107301 and NIDDK R01DK121072 and 1RO3DK117252. R.E.S. is supported as Irma Hirschl Trust Research Award Scholar. We would like to thank Englander Institute of Precision Medicine Mass Cytometry Core facility for processing, labeling, and acquiring samples for IMC imaging. We thank Zenodo for storing the raw data associated with this manuscript with a higher usage quota.

## Author contributions

A.F.R., H.R., A.B., O.E., and R.E.S. planned the study; A.B., Y.B. and S.S. provided and evaluated samples and clinical data; H.R. performed imaging mass cytometry; A.F.R., performed analysis of the data. O.E., and R.E.S. supervised the research. A.F.R., H.R., A.B., O.E., and R.E.S. wrote the manuscript with contributions from all authors.

## Competing financial interests

O.E. is scientific advisor and equity holder in Freenome, Owkin, Volastra Therapeutics and OneThree Biotech. R.E.S. is a member of the scientific advisory board of Miromatrix Inc. The remaining authors declare no competing financial interests.

## Notes

### Author Declarations

The Tissue Procurement Facility operates under Institutional Review Board (IRB) approved protocol and follows guidelines set by Health Insurance Portability and Accountability Act (HIPAA). Experiments using samples from human subjects were conducted in accordance with local regulations and with the approval of the IRB at the Weill Cornell Medicine. The autopsy samples were collected under protocol 20-04021814.

