## Extended data for "The spatio-temporal landscape of lung pathology in SARS-CoV-2 infection"

[1] Institute for Computational Biomedicine, Weill Cornell Medicine, New York, NY, USA
[2] Caryl and Israel Englander Institute for Precision Medicine, Weill Cornell Medicine, New York, NY, USA
[3] WorldQuant Initiative for Quantitative Prediction
[4] Department of Medicine, Weill Cornell Medicine, New York, NY, USA
[5] Department of Pathology and Laboratory Medicine, Weill Cornell Medicine, New York, NY, USA
[*] Co-first authors
[Ω] Co-senior authors

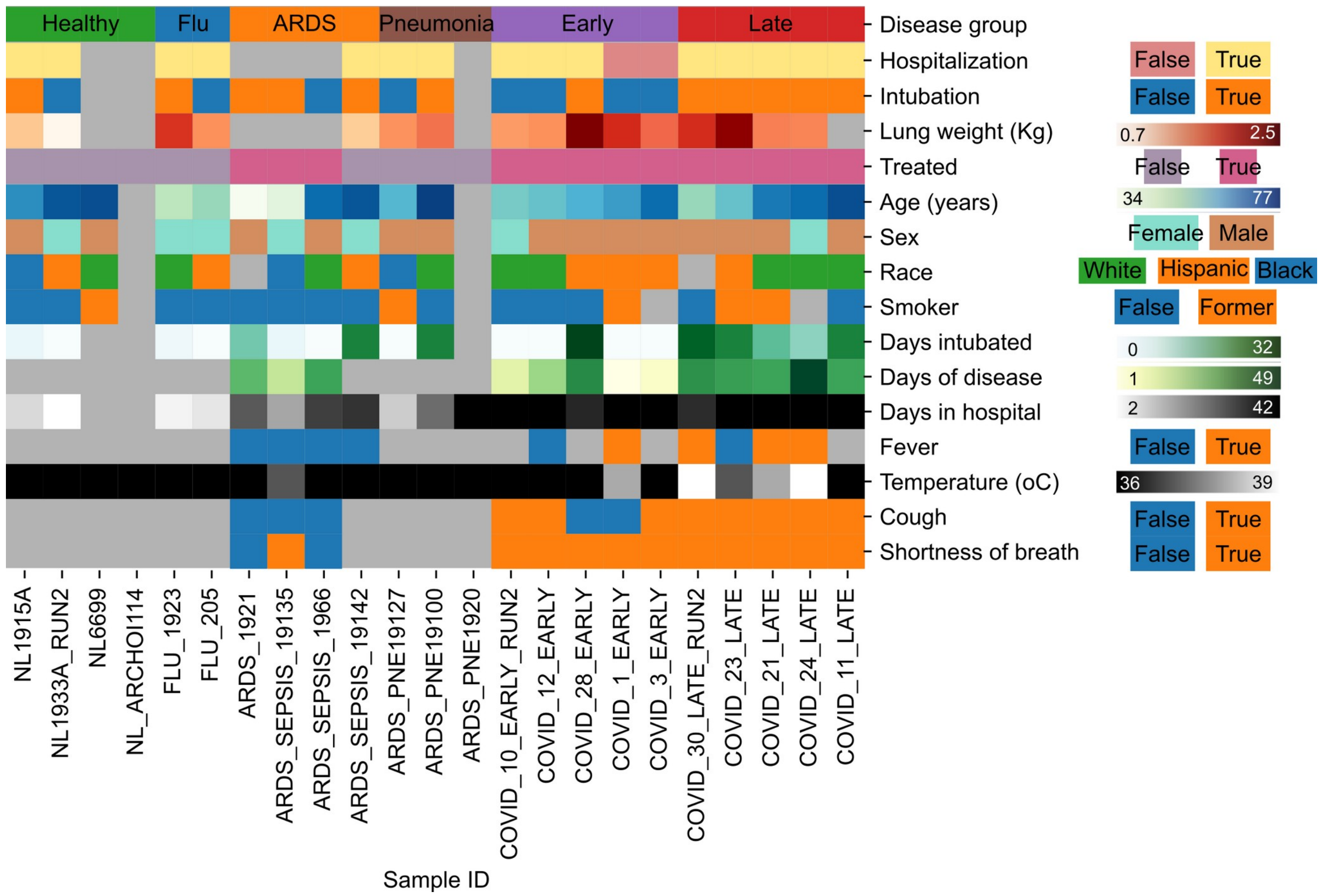


**Extended Data 1:** Clinical and demographic information for patients in the cohort.
Heatmap depicting the values of each individual for all acquired clinical and demographic variables. Grey color indicates missing or non-applicable values.

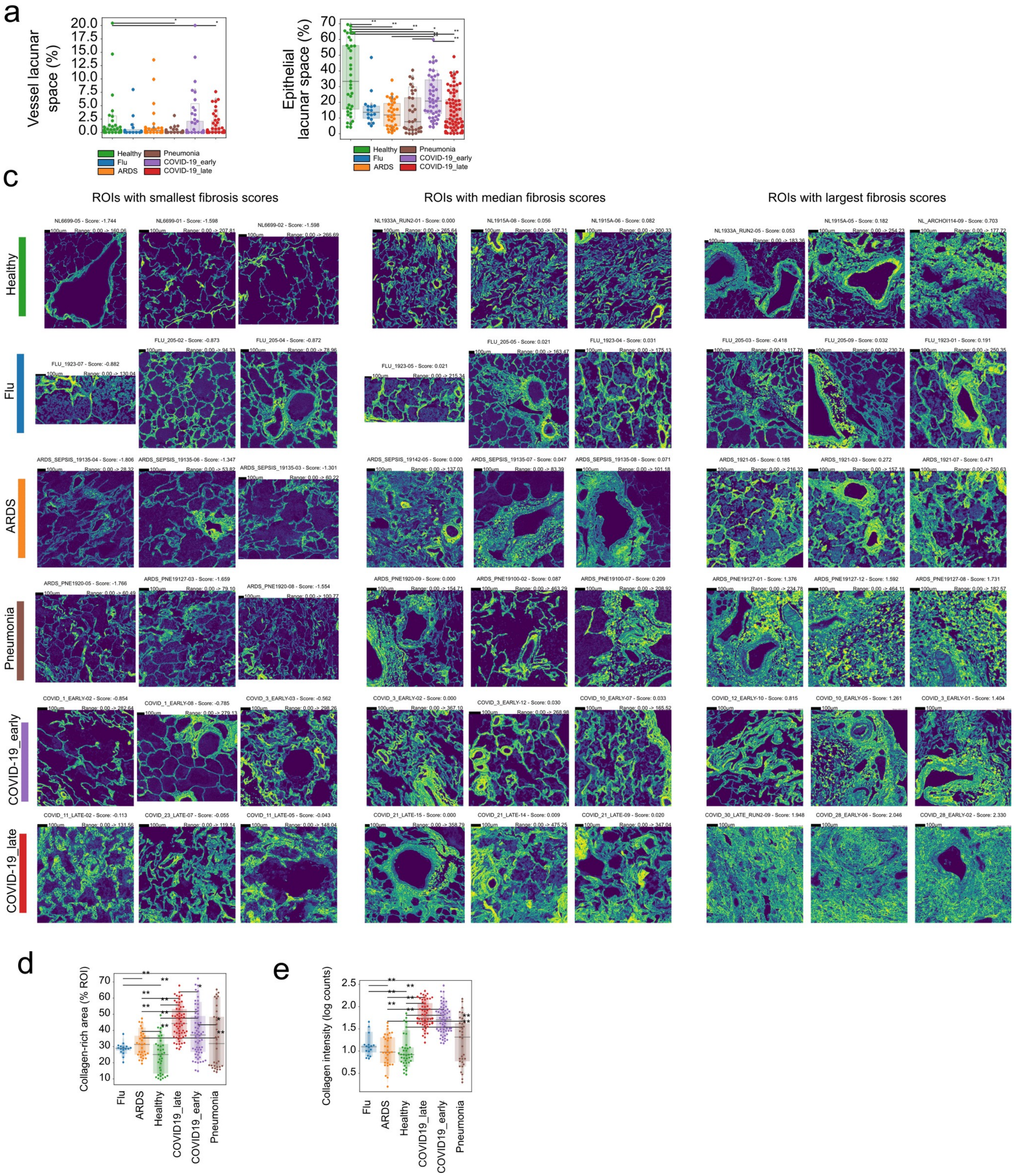

**Extended Data 2:** Quantification of macroscopic presentation of lung tissue.
**a-b)** Percentage of lacunar space attributed to a) vessel or b) epithelial space per image grouped by disease. **c)** Collagen type I in images from lungs of healthy individuals, or lung pathology patients and the associated fibrosis score. Images with lowest, median and highest fibrosis scores are depicted. **d)** Percentage of image covered in Collagen type I for each image grouped by disease group. **e)** Mean intensity of Collagen type I in lung IMC images grouped by disease group.

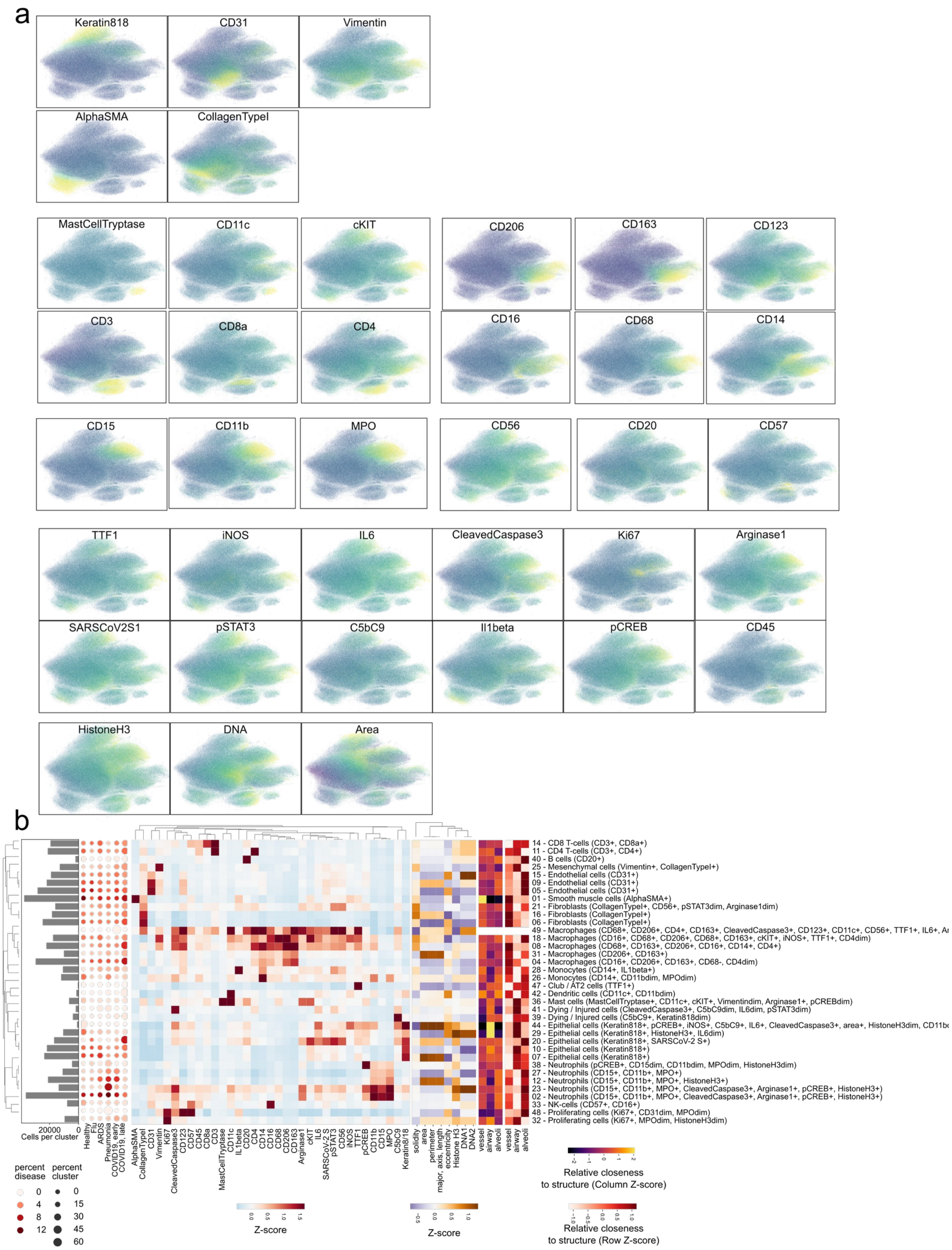


**Extended Data 3:** Unsupervised cell type identification.

**a)** UMAP projection of all single-cells where cells are colored by the intensity of each channel. **b)** Hierarchically clustered heatmap of discovered clusters (rows) and the mean intensity of each channel (columns) for each. The histogram on the left represents the absolute abundance of each cluster across all images. The dot-plot represents the relative abundance of each cluster in each disease group.

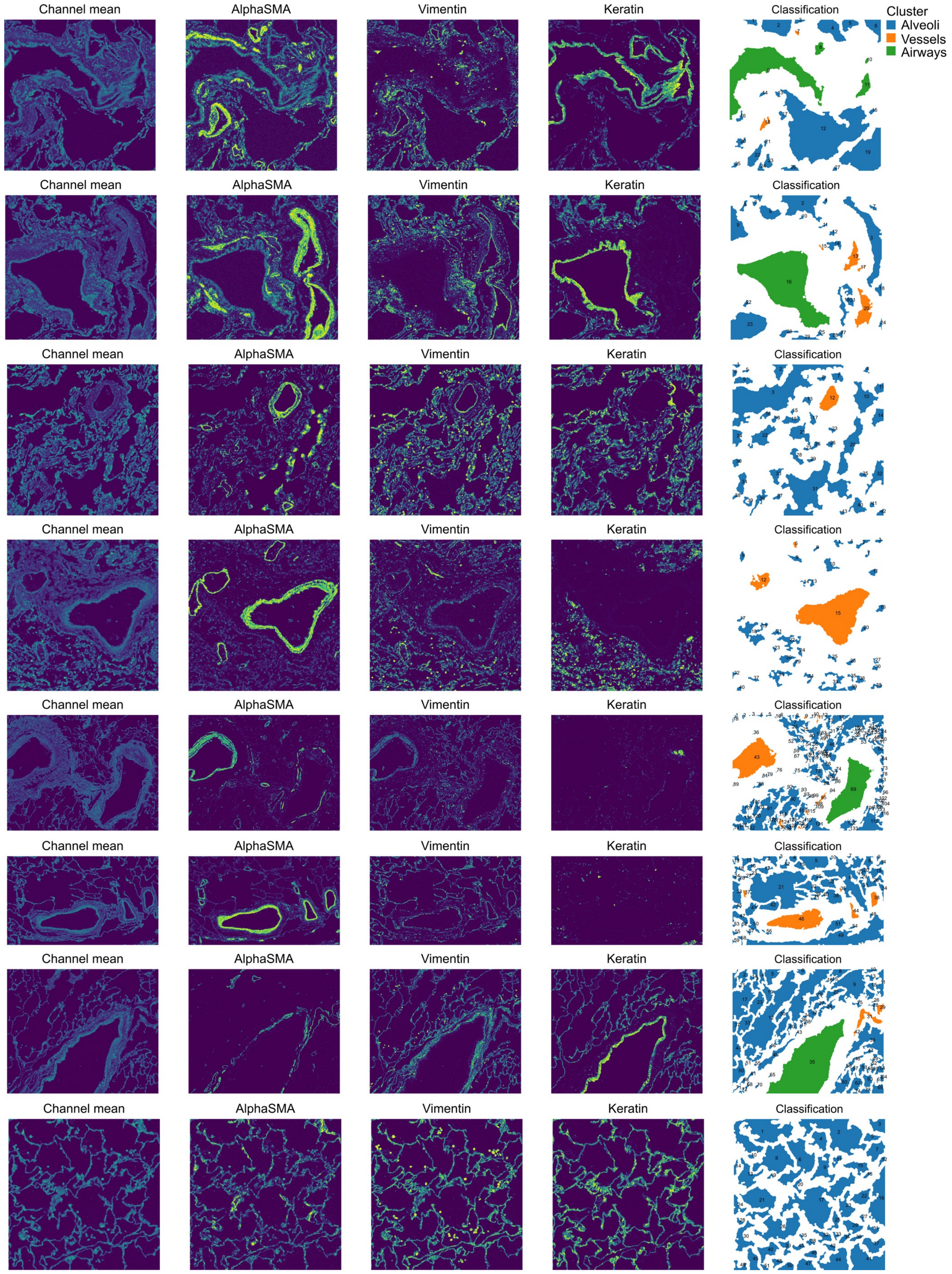
Channel mean
AlphaSMA
Vimentin
Keratin
Classification
Cluster
Alveoli
Vessels
Airways

**Extended Data 4:** Classification of lung lacunae.
Representative images of healthy lung images with the mean of all channels and channels important to discern between vessels, airways and alveoli. The last column represents the final classification of lacunae into each of the three classes of structures.

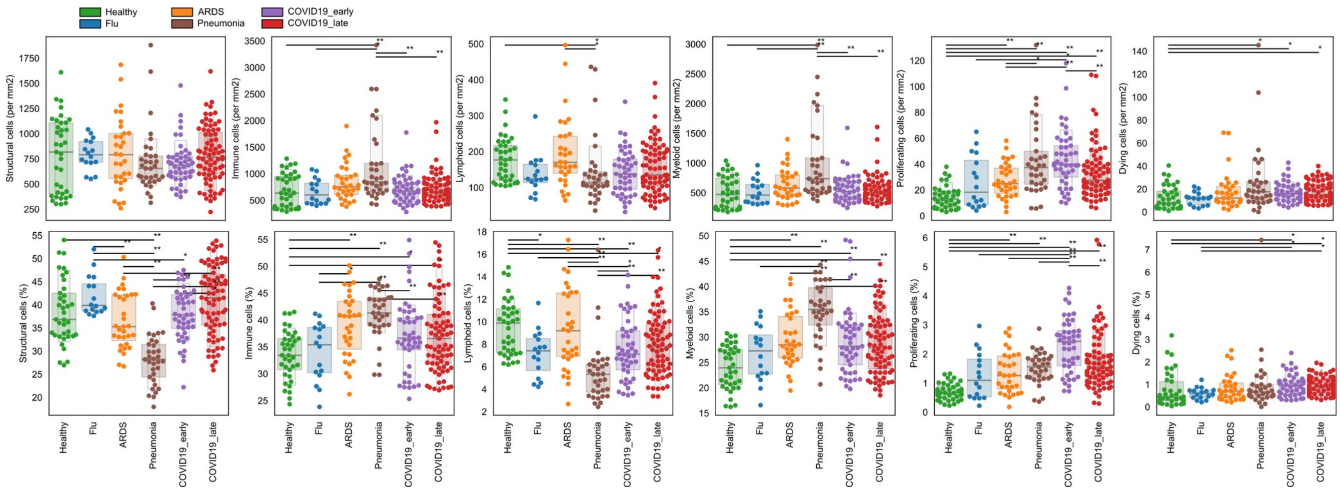


**Extended Data 5:** Global abundance of structural and immune cells.
Absolute (first row) and relative (second row) abundance of groups of cells dependent on disease group.

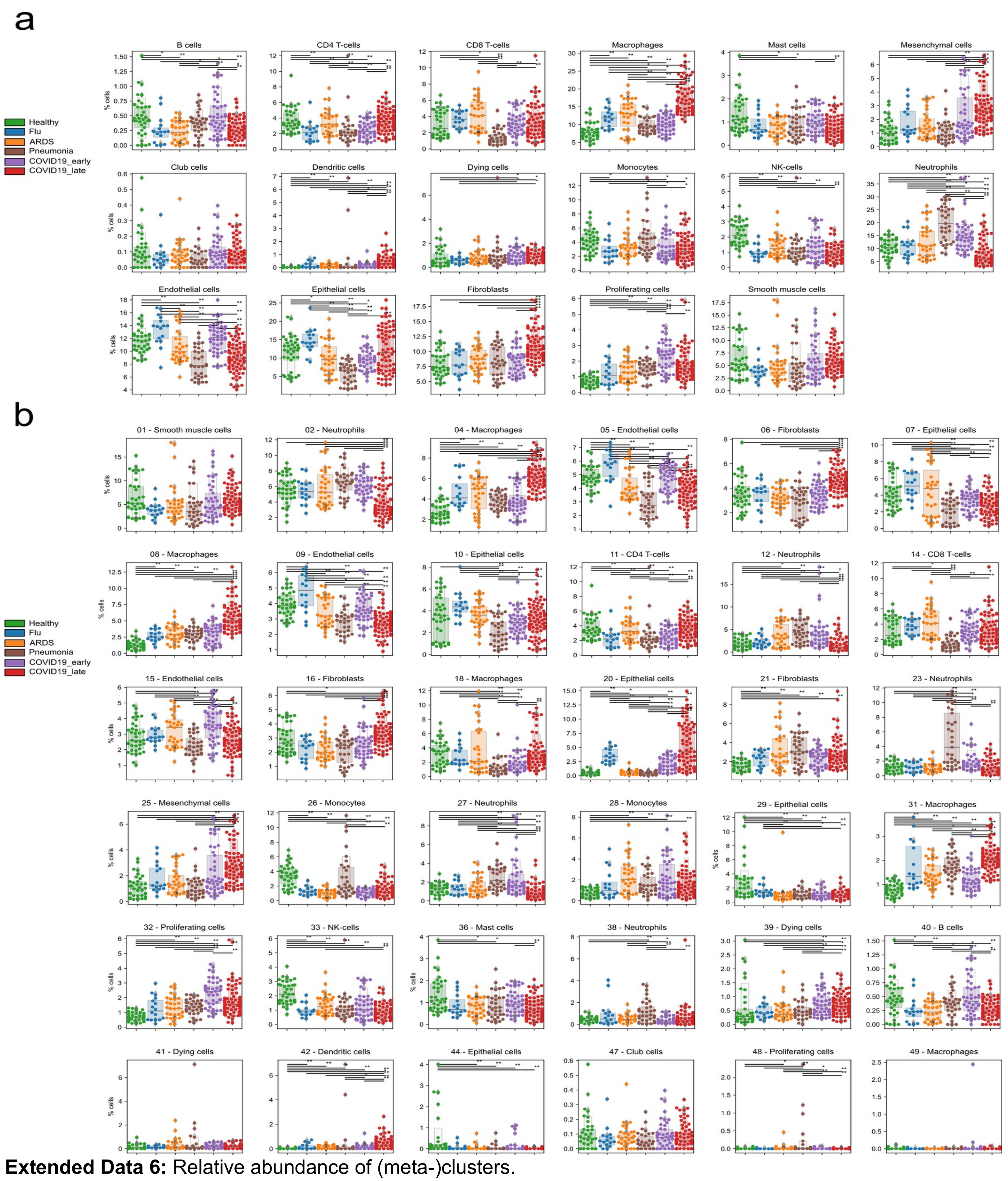


**Extended Data 6:** Relative abundance of (meta-)clusters.
**a-b)** Relative abundance of a) meta-clusters or b) clusters as fraction of total cells per image, grouped by disease group.

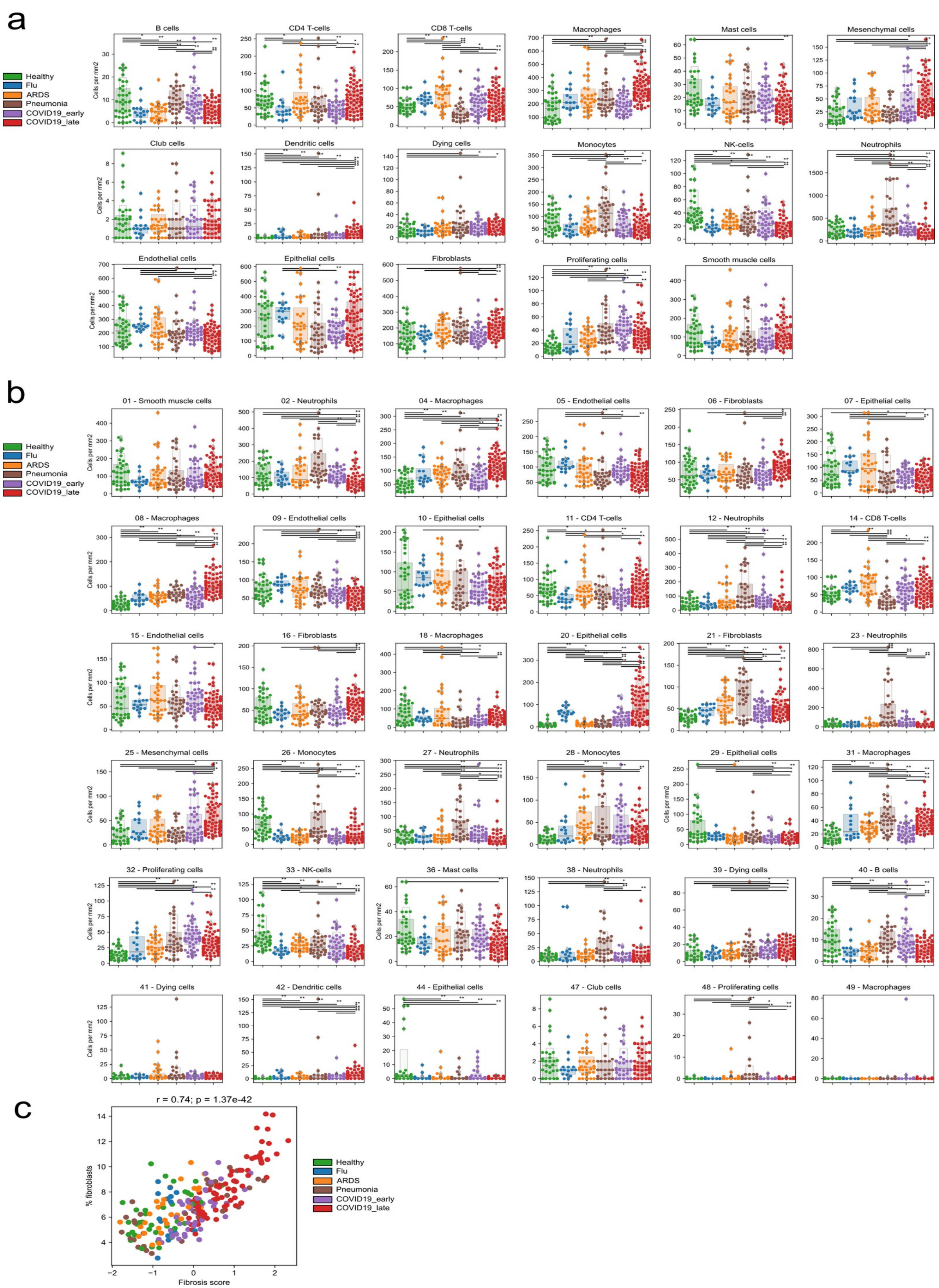


**Extended Data 7:** Absolute abundance of (meta-)clusters.
**a-b)** Absolute abundance of a) meta-clusters or b) clusters per image, grouped by disease group. **c)** Relationship between fibrosis score and fibroblast meta-cluster abundance visualized as a scatter plot.

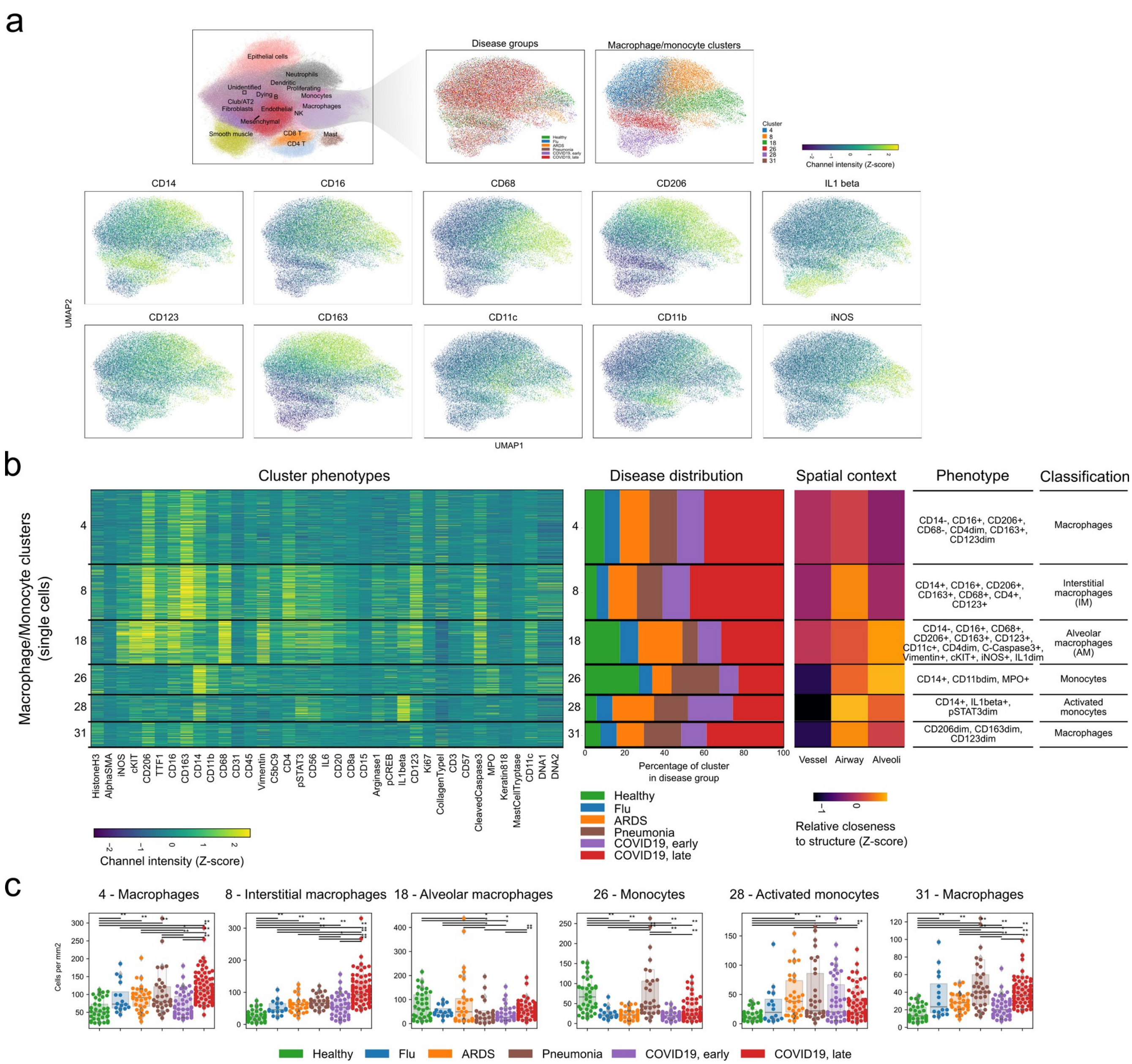


**Extended Data 8:** Diversity of myeloid cells.

**a)** UMAP representation of myeloid cells and the prominent markers associated with them. **b)** Phenotypic markers, spatial context and abundance in disease groups for each of the 6 myeloid clusters. **c)** Abundance of each myeloid cluster in the disease groups. Each point represents the abundance of that cluster in a given region of interest.

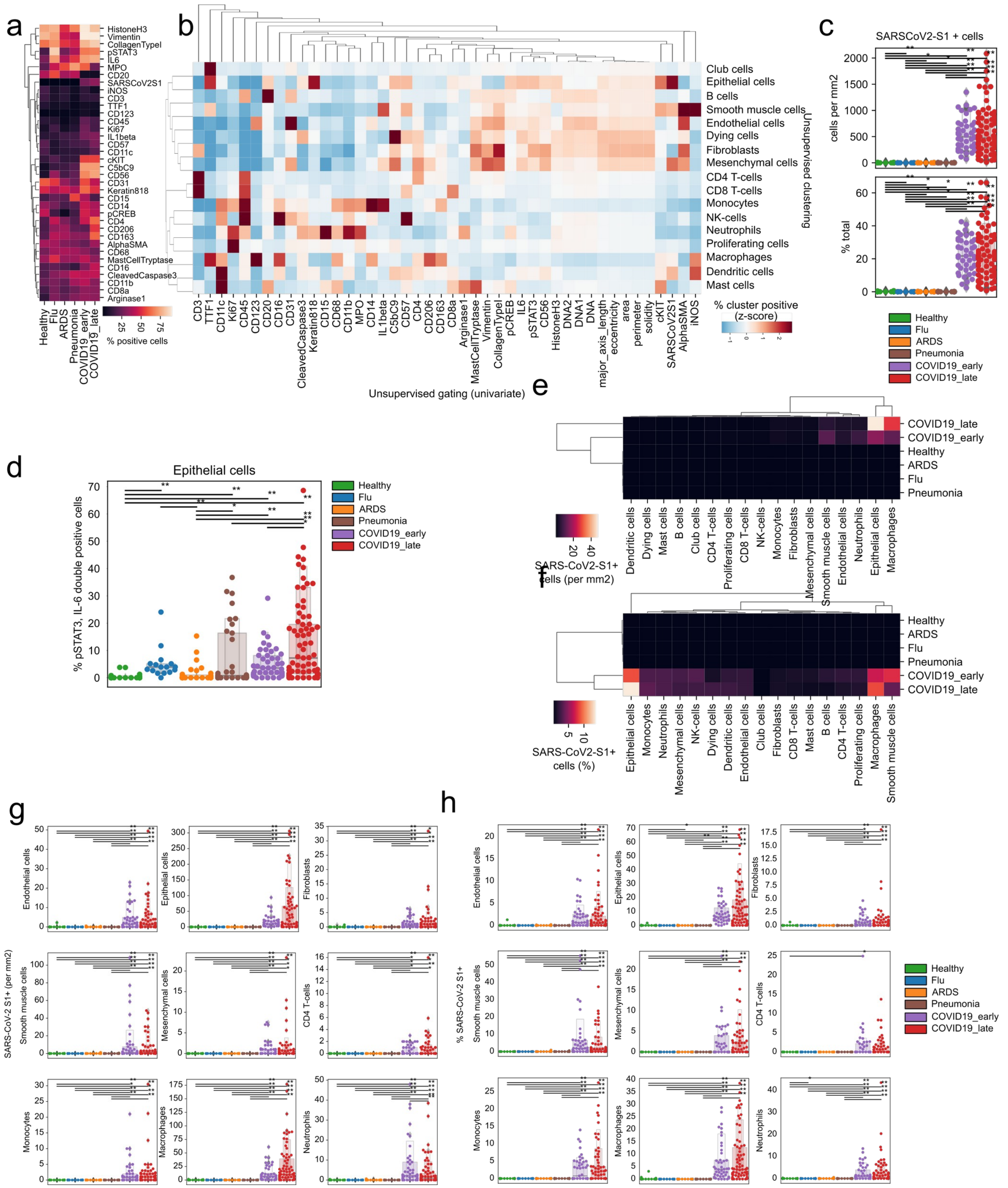


**Extended Data 9:** Identification of cell states.

**a)** Percentage of cells positive for each IMC channel as classified by univariate Gaussian mixture models per disease group. **b)** Percentage of channel positive cells per each meta-cluster. Values represent a column-wise Z-score. **c)** Absolute (top) and relative (bottom) frequency of SARS-CoV-2 Spike$^{+}$ cells per disease group. **d-e)** d) Absolute and e) proportional amount of SARS-CoV-2 Spike$^{+}$ cells grouped per meta-cluster and disease group. **f-g)** f) Absolute and g) proportional frequencies of cells positive for SARS-CoV-2 Spike$^{+}$ cells per meta-cluster and disease group.

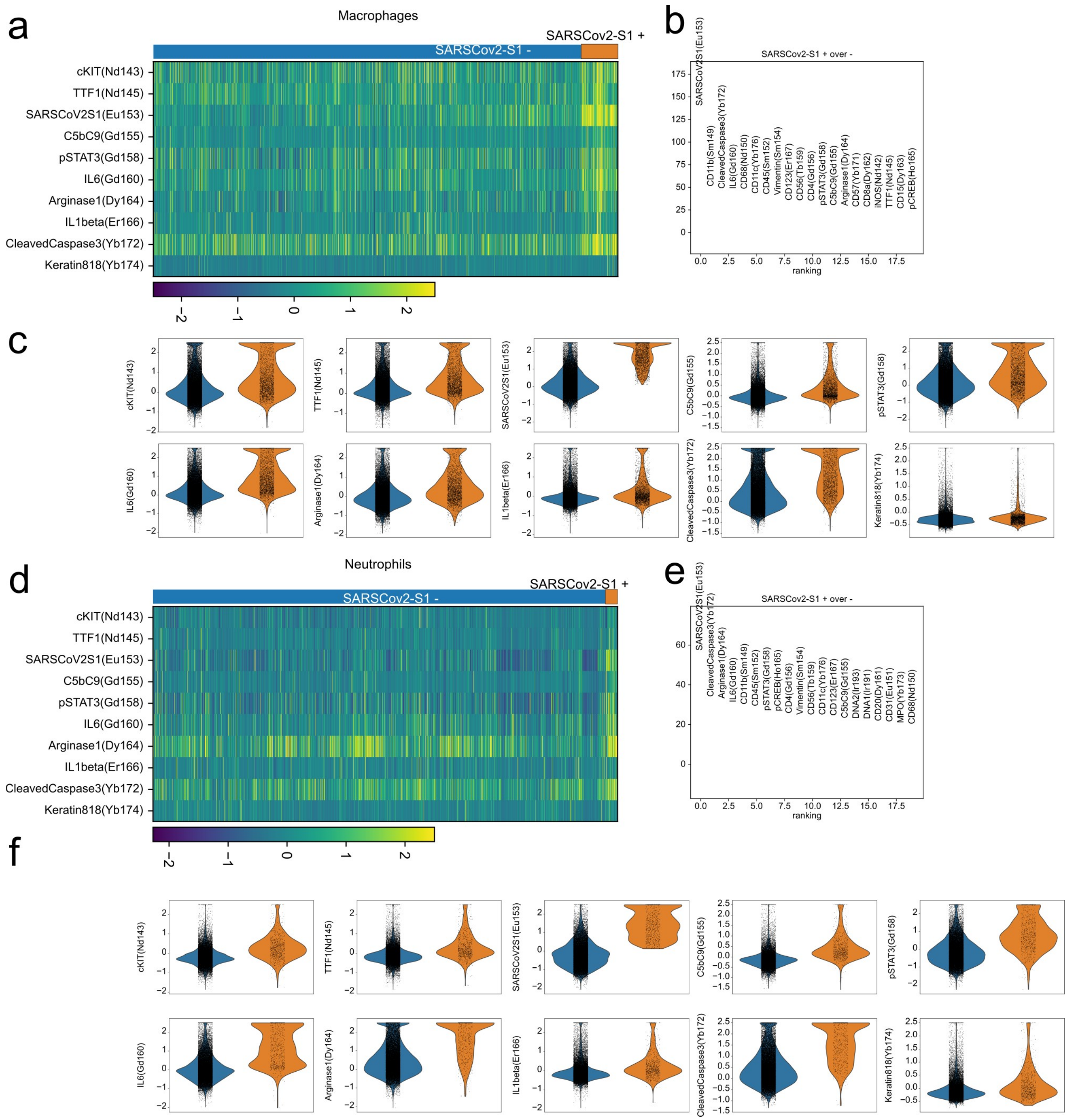


**Extended Data 10:** Phenotype of SARS-CoV-2 Spike⁺ cells.
**a)** Heatmap of single macrophage cells (columns) and functional markers (rows) with cells grouped by SARS-CoV-2 Spike positivity. **b)** Score of differential expression for IMC channels dependent on SARS-CoV-2 Spike positivity status of macrophage cells. **c)** Intensity of IMC channels per single-cell dependent on SARS-CoV-2 Spike positivity. **d)** Heatmap of single neutrophil cells (columns) and functional markers (rows) with cells grouped by SARS-CoV-2 Spike positivity. **e)** Score of differential expression for IMC channels dependent on SARS-CoV-2 Spike positivity status of neutrophils cells. **f)** Intensity of IMC channels per single-cell dependent on SARS-CoV-2 Spike positivity.

a

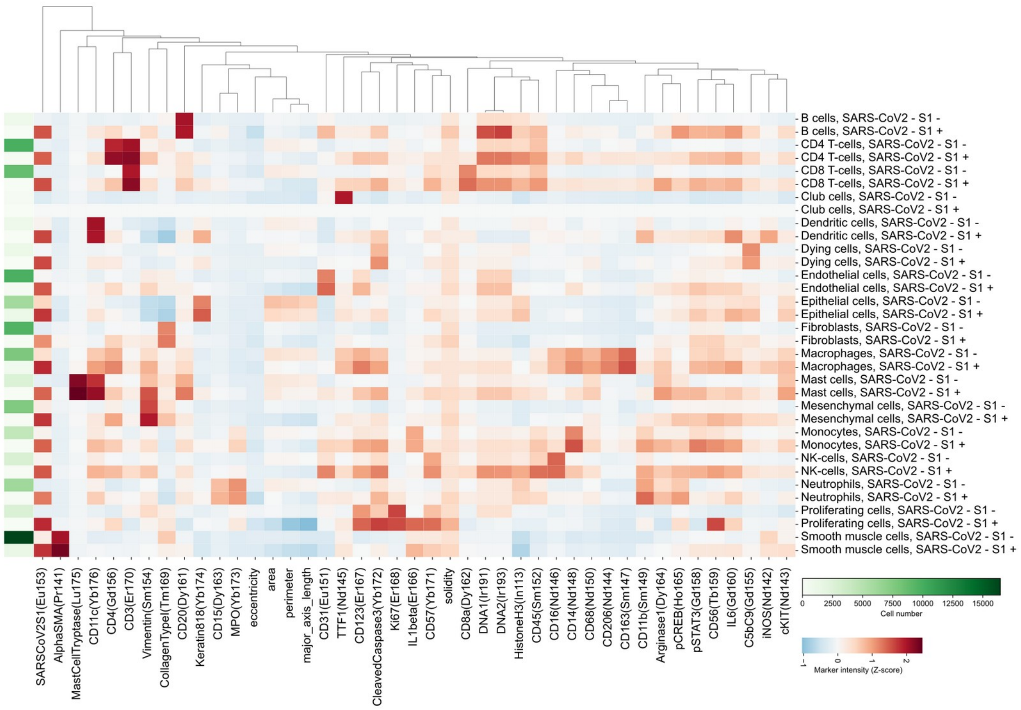


b

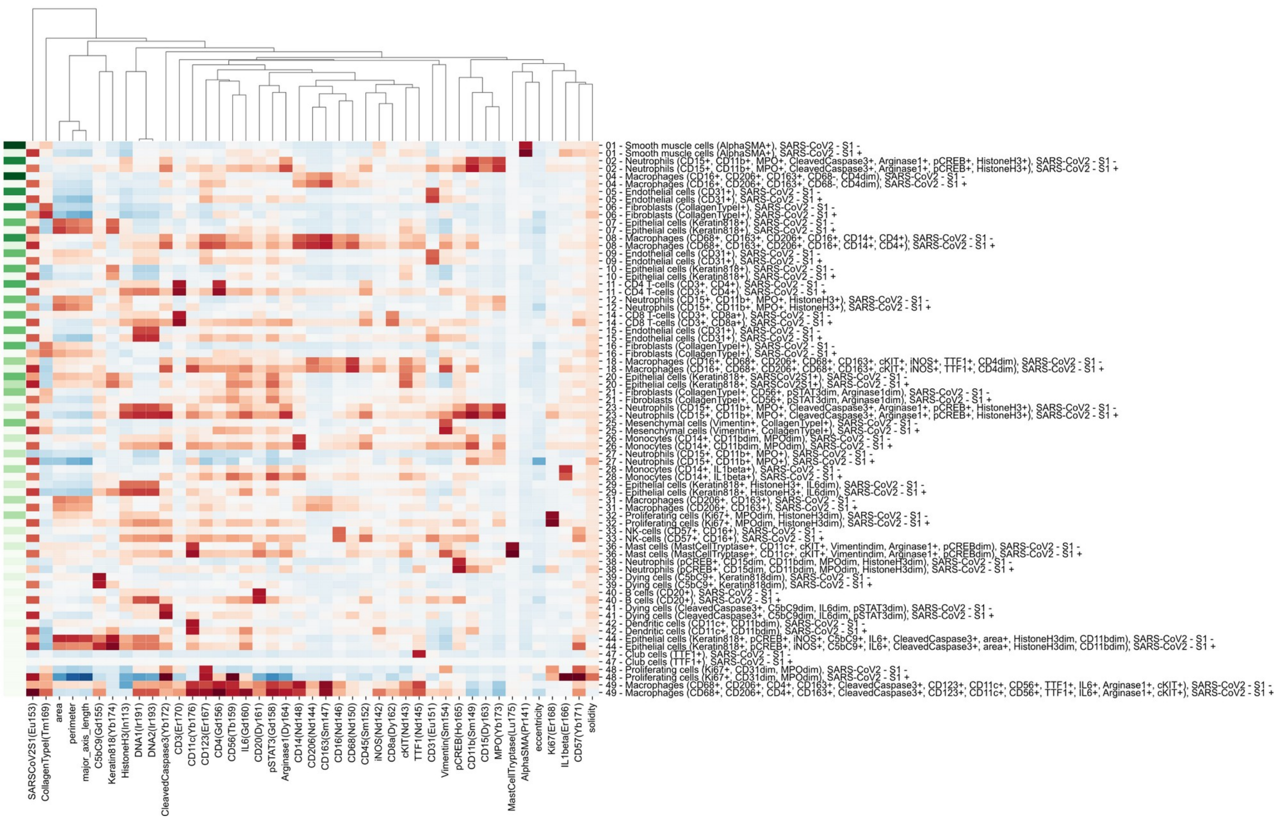

**Extended Data 11:** Phenotype of SARS-CoV-2 Spike⁺ cells for all cell types.
**a-b)** Mean IMC channel intensity for all a) meta-clusters or b) clusters dependent on SARS-CoV-2 Spike positivity.

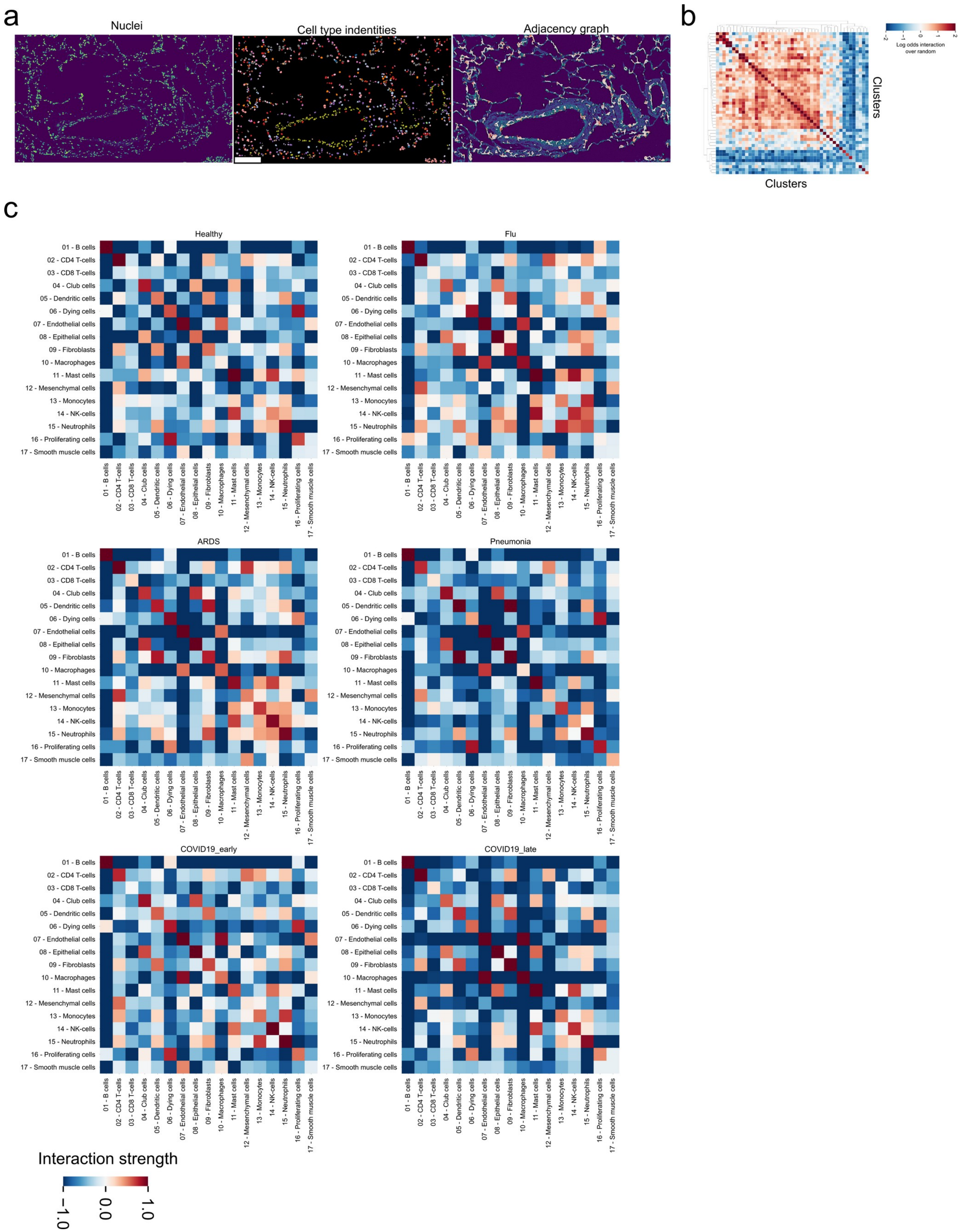


**Extended Data 12:** Cellular interactions between cell types in COVID-19.
**a)** Exemplary description of the derivation of a Region Adjacency Graph (RAG) for a given lung IMC image. The leftmost image depicts the DNA channel marking nuclei, the centermost the identified meta-clusters, and the

rightmost the RAG represented as edges between adjacent cells. **b)** Observed values of pairwise cluster interactions over the expected values for the same cellular interactions for the image in a). **c)** Pairwise interactions between meta-clusters aggregated by the mean value across images depending on the disease group.

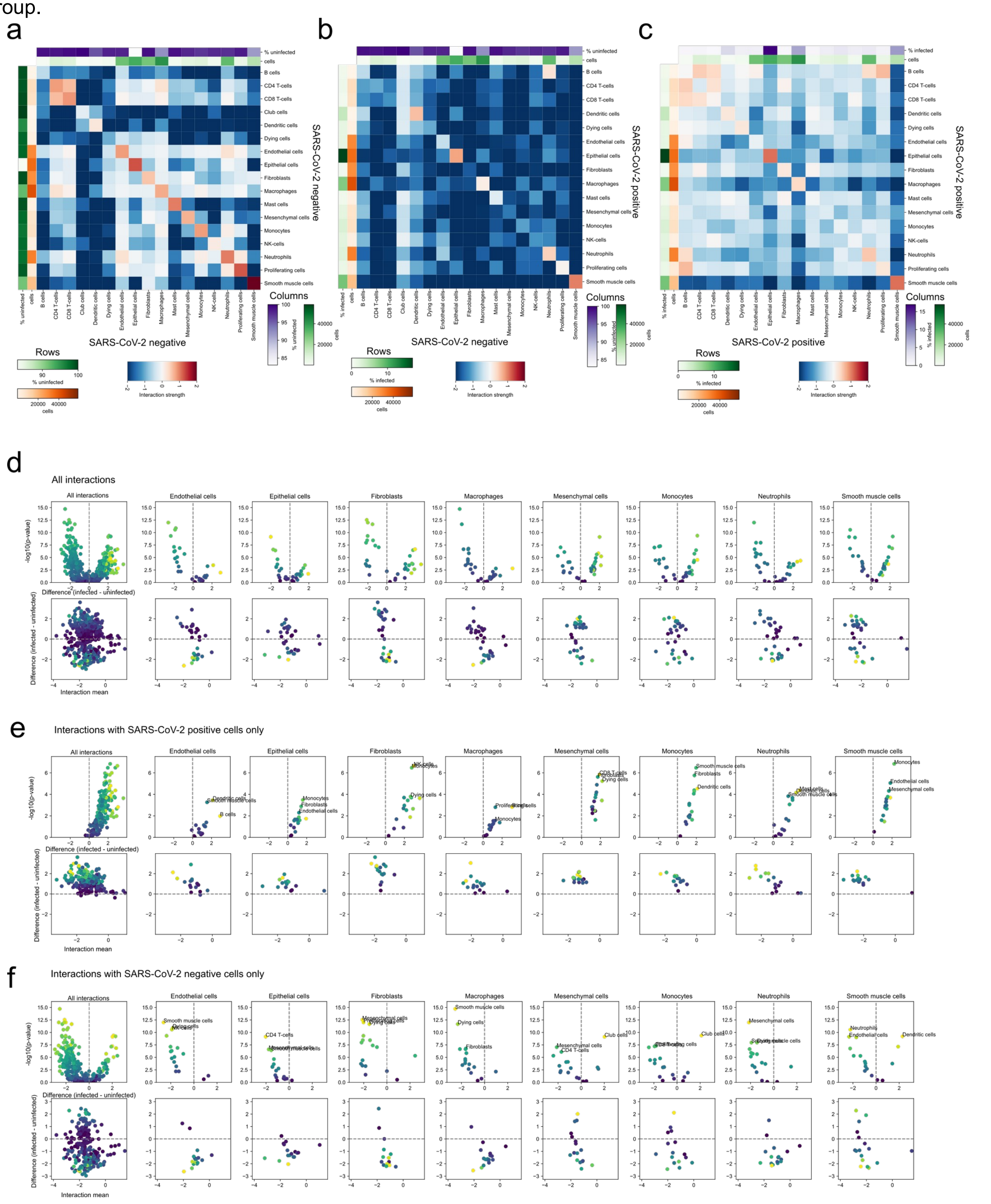


**Extended Data 13:** Effect of SARS-CoV-2 Spike⁺ on COVID-19 cellular interactions.
**a-c)** Pairwise cellular interactions between meta-clusters dependent on SARS-CoV-2 Spike positivity: a) uninfected cells; b) between SARS-CoV-2 Spike positive and negative cells; c) between infected cells. **d-f)**

Statistical testing of differential interactions of infected cells and other cell types and uninfected cells and other cell types, dependent on the SARS-CoV-2 Spike positivity of the second cell type: d) both SARS-CoV-2 Spike$^-$ and SARS-CoV-2 Spike$^+$ cells; e) only SARS-CoV-2 Spike$^+$ cells; f) only SARS-CoV-2 Spike$^-$ cells. The top rows display a volcano plot (x-axis: difference in interaction between SARS-CoV-2 Spike$^+$ and SARS-CoV-2 Spike$^-$ cells; y-axis: -log10 Mann-Whitney U-test FDR-adjusted p-value), while the bottom rows display an MA-plot (x-axis: mean of interaction score across all images; y-axis: difference in interaction between SARS-CoV-2 Spike$^+$ and SARS-CoV-2 Spike$^-$ cells).

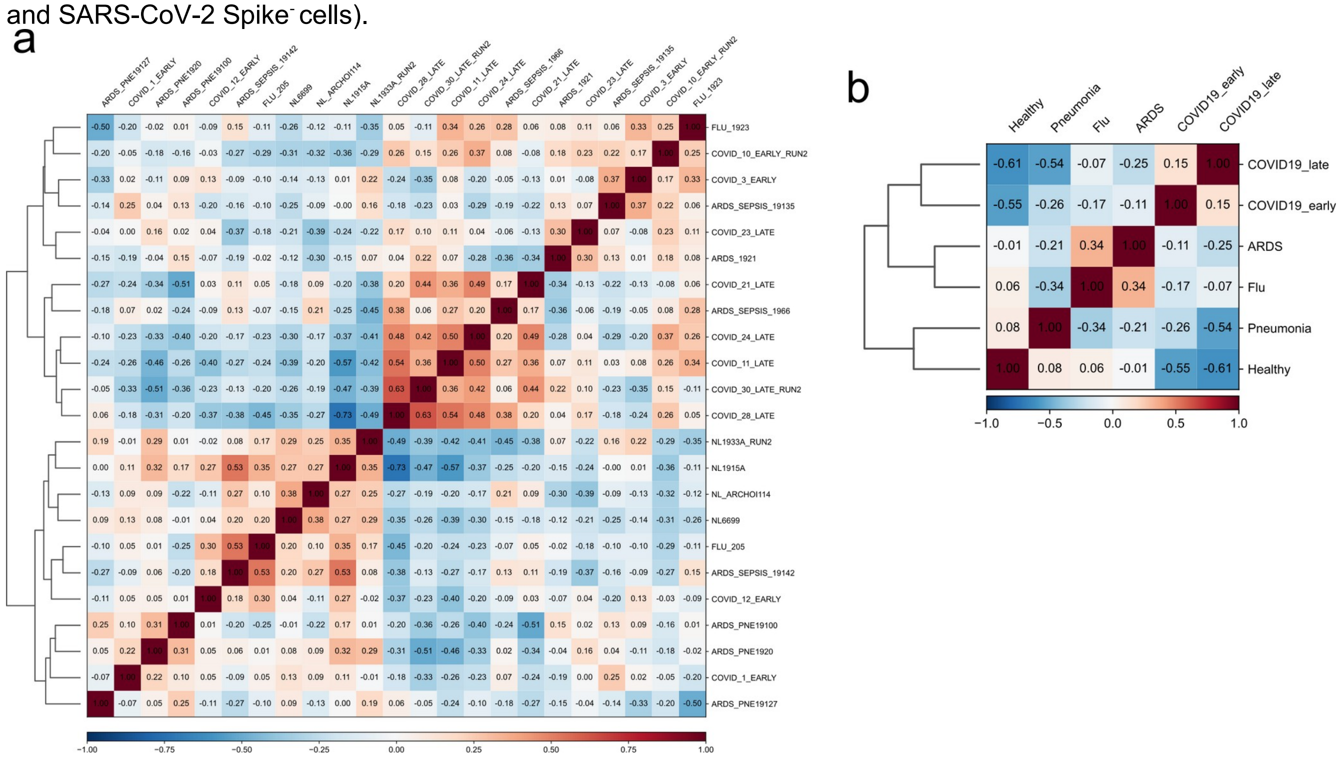


**Extended Data 14:** Relationship between IMC samples and disease groups.
**a-b)** Pairwise Pearson correlation of cell type abundances between a) IMC samples b) disease groups.

a

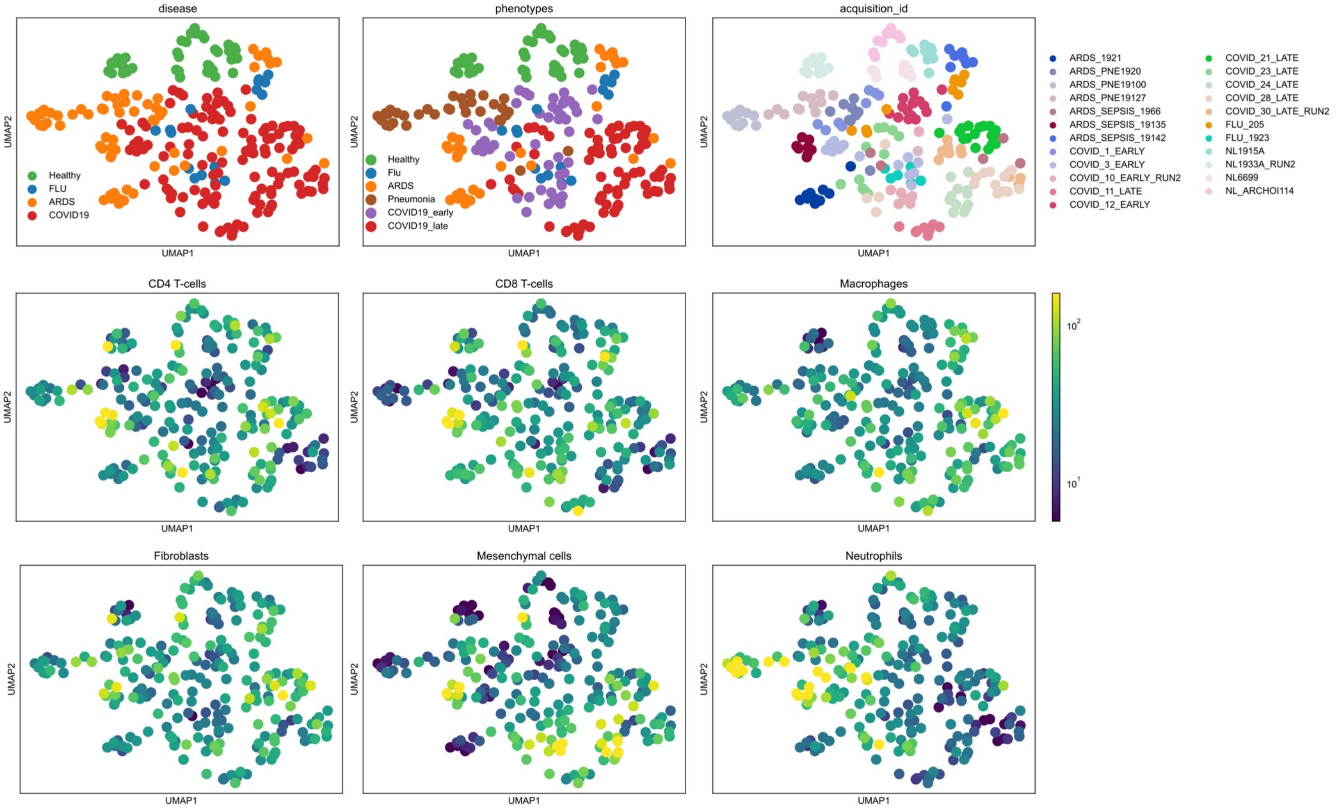


b

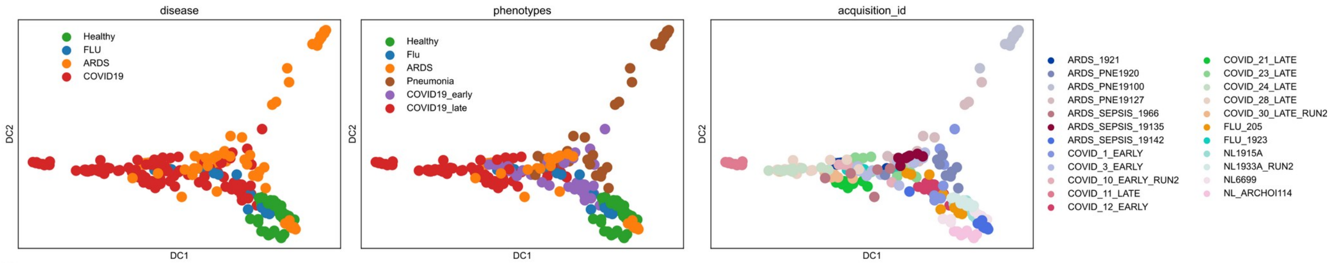


c

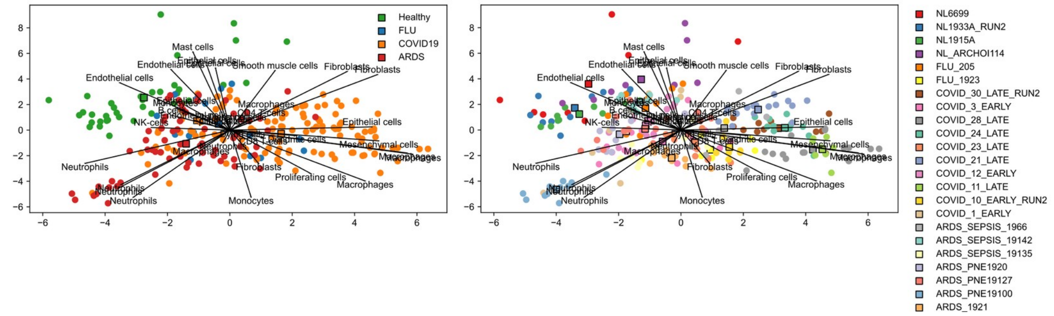


**Extended Data 15:** Alternative methods to derive an unsupervised pathology landscape.
**a-c)** a) UMAP, b) Diffusion map or c) PCA projection of IMC images colored by disease group, subgroup or sample ID.

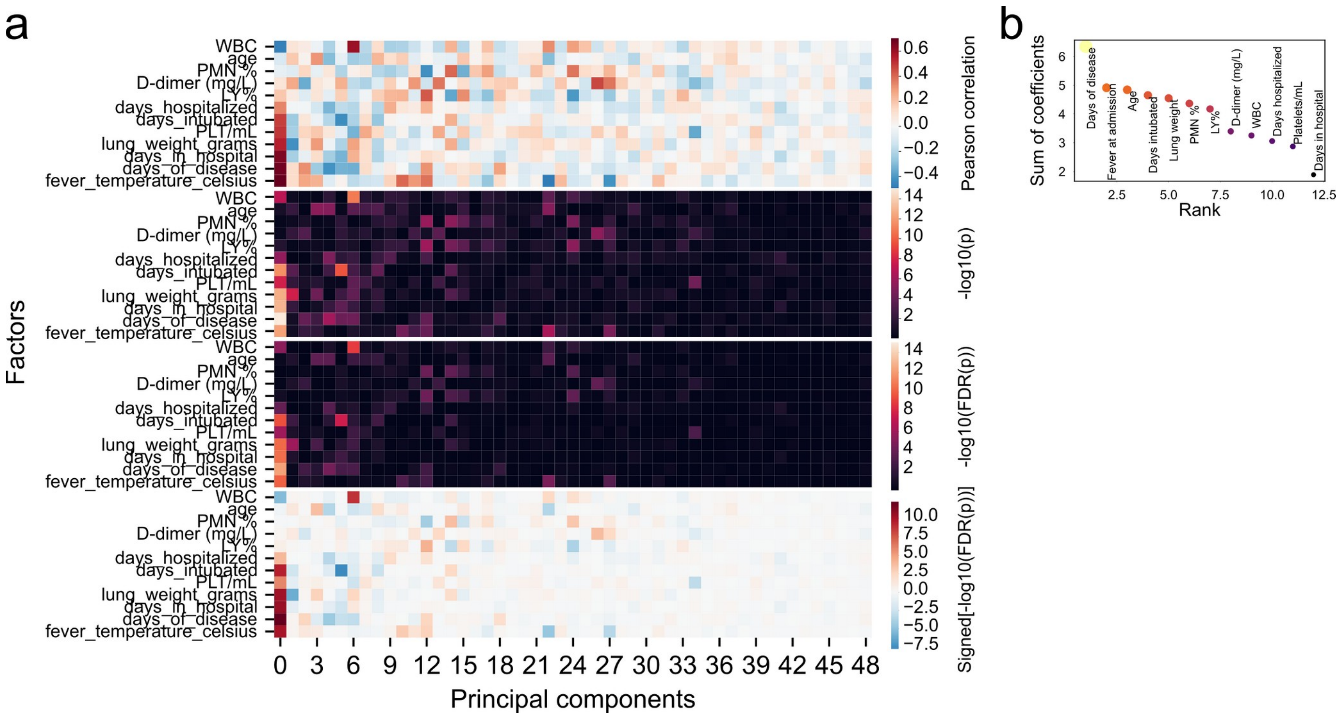


**Extended Data 16:** Association analysis of clinical factors and principal components.
**a)** Correlation coefficients (top) or p-values of association between clinical factors and principal components. **b)** Sum of absolute correlation coefficients across all principal components.

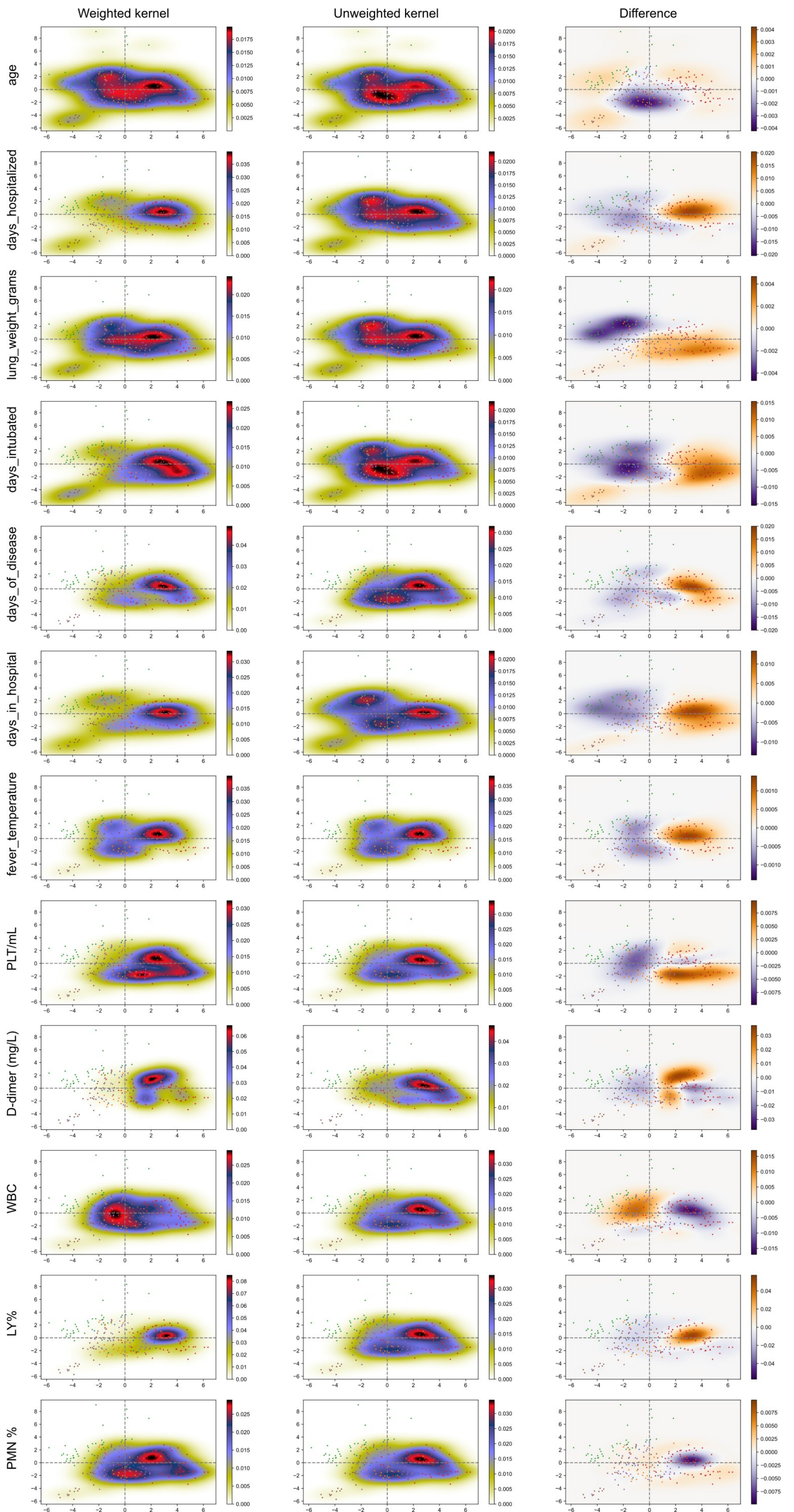
Weighted kernel
Unweighted kernel
Difference
age
days_hospitalized
lung_weight_grams
days_intubated
days_of_disease
days_in_hospital
fever_temperature
PLT/mL
D-dimer (mg/L)
WBC
LY%
PMN %

**Extended Data 17:** Projection of clinical factors onto pathology landscape.
**a-c)** Kernel density estimation for various clinical and demographic factors a) weighted by the factor values, b) unweighted, or c) the difference of a) and b).

### Supplementary table legends

**Supplementary Table 1**: Clinical and demographic information for patients in the cohort.

**Supplementary Table 2**: Description of antibodies used in the study.

**Supplementary Table 3**: Statistical testing of difference in abundance for clusters and meta-clusters between disease groups.

**Supplementary Table 4**: Statistical testing of difference in fraction of cells positive in functional markers for clusters and meta-clusters between disease groups.
